# Health-economic burden attributable to novel serotypes in candidate 24- and 31-valent pneumococcal conjugate vaccines

**DOI:** 10.1101/2024.04.11.24305684

**Authors:** Laura M King, Joseph A Lewnard

## Abstract

**Background:** Next-generation pneumococcal vaccines currently in clinical trials include 24- and 31-valent pneumococcal conjugate vaccines (PCV24, PCV31), which aim to prevent upper-respiratory carriage and disease involving the targeted serotypes. We aimed to estimate the comprehensive health-economic burden associated with acute respiratory infections (ARIs) and invasive pneumococcal disease (IPD) attributable to PCV24- and PCV31-additional (non-PCV20) serotypes in the United States.

**Methods:** We multiplied all-cause incidence rate estimates for acute otitis media (AOM), sinusitis, and non-bacteremic pneumonia by estimates of the proportions of each of these conditions attributable to pneumococci and the proportions of pneumococcal infections involving PCV24- and PCV31-additional serotypes. We obtained serotype-specific IPD incidence rates from US Active Bacterial Core surveillance data. We accounted for direct medical and non-medical costs associated with each condition to estimate resulting health-economic burden. Non-medical costs included missed work and lost quality-adjusted life years due to death and disability.

**Results:** The health-economic burden of PCV24-additional serotypes totaled $1.3 billion ($1.1-1.7b) annually in medical and non-medical costs, comprised of $0.9b ($0.7-1.2b) due to ARIs and $0.4b ($0.3-0.5b) due to IPD. For PCV31-additional serotypes, medical and non-medical costs totaled $7.5b ($6.6-8.6b) annually, with $5.5b ($4.7-6.6b) due to ARIs and $1.9b ($1.8-2.1b) due to IPD. The largest single driver of costs was non-bacteremic pneumonia, particularly in adults aged 50-64 and ≥65 years.

**Conclusions:** Additional serotypes in PCV24 and PCV31, especially those included in PCV31, account for substantial health-economic burden in the United States.

**Summary:** Novel serotypes in 24- and 31-valent pneumococcal conjugate vaccines, currently in clinical trials, account for substantial health-economic burdens in the United States, especially in adults ≥50 years. Vaccines targeting these serotypes may prevent pneumococcal disease and reduce associated costs.

## BACKGROUND

*Streptococcus pneumoniae* (pneumococcus) causes disease ranging from mild, outpatient-managed acute respiratory infections (ARIs) such as acute otitis media (AOM) and sinusitis to severe pneumonia and invasive infections (e.g., meningitis, bacteremia). Pneumococcus accounts for an estimated 19-46% of outpatient pediatric ARIs,^1^ and invasive pneumococcal disease (IPD) incidence totaled 9.2 cases per 100,000 population in 2019.^2^

Pneumococcal conjugate vaccines (PCVs) reduce upper respiratory carriage of vaccine-type *S. pneumoniae* serotypes and their progression to disease,^3,4^ making PCVs important public health strategies for mitigating pneumococcal disease burden. Implementation of previous (e.g., 7- and 13-valent) PCV products was found to be cost-saving due to substantial reductions in pneumococcal disease-associated healthcare and societal expenditures.^5,6^ However, selective pressure from PCV-conferred immunity has resulted in increases in disease caused by non-vaccine type serotypes—a phenomenon known as serotype replacement.^7^ Thus, next-generation PCVs with additional serotypes must be considered continually. Beginning in 2023, 15- and 20-valent PCVs (PCV15, PCV20) were recommended for routine use in infants in a schedule of 3 primary doses at 2,4, and 6 months and a booster between 12-15 months of age.^8,9^ Additionally, PCV-naïve adults aged ≥65 years and those aged <65 years with high-risk comorbid conditions were recommended to receive either PCV20 alone or PCV15 in series with 23-valent pneumococcal polysaccharide vaccine (PPSV23).^10^

Two candidate next-generation pneumococcal vaccines currently in clinical trials include 24- and 31-valent PCVs (PCV24, PCV31) employing a novel conjugation approach involving the CRM197 carrier protein.^11^ These formulations expand upon the serotypes included in PCV20 (1, 3, 4, 5, 6A, 6B, 7F, 8, 9V, 10A, 11A, 12F, 14, 15B, 18C, 19A, 19F, 22F, 23F, X33F) to include serotypes 2, 9N, 17F, 20B (PCV24/PCV31) and 7C, 15A, 16F, 23A, 23B, 31, 35B (PCV31). However, burden associated with PCV24- and PCV31-additional (non-PCV20) serotypes has not been quantified. We aimed to estimate the comprehensive health-economic burden attributable to ARIs and IPD caused by PCV24- and PCV31-additional serotypes in the United States.

## METHODS

### Non-invasive disease incidence and etiology

We used a three-component approach to estimate outpatient visits, antibiotic prescriptions, and hospitalizations due to AOM, sinusitis, non-bacteremic pneumonia, and mastoiditis (a complication of AOM) caused by PCV24- and PCV31-additional serotypes. We multiplied all-cause incidence rates for each outcome by 1) estimates of the proportion of cases caused by pneumococcus, and 2) estimates of the proportion of all pneumococcal isolates associated with PCV24- and PCV31-additional serotypes for these conditions. To estimate absolute case volumes as of 2019 (selected as the most recent year indicating conditions without disruption of healthcare delivery patterns and pneumococcal disease dynamics due to the COVID-19 pandemic), we multiplied the resulting age-specific incidence rate estimates by corresponding 2019 Bridged-Race census population estimates.^12^

We fit gamma distributions to reported all-cause incidence rate estimates for each condition from published studies (**Tables S1-S2**) and drew from the resulting distributions to propagate uncertainty in incidence rates. In identifying studies for inclusion, we prioritized recent, nationally-representative studies, where available. We similarly used data from published studies of pneumococcal-attributable proportions of cases for each condition (**Table S3**), sampling from beta distributions to propagate uncertainty.

To estimate the proportion of pneumococcal infections involving PCV24- and PCV31-additional serotypes, we conducted meta-analyses of studies describing serotype distributions among pneumococcal isolates collected from individuals experiencing each condition (**Table 1**). Defining counts of serotype-specific isolates from each study as draws from a multinomial distribution, we used Markov-chain Monte Carlo to sample underlying serotype distributions among pneumococcal infections, handling serotype-specific proportions for each condition as a Dirichlet parameter vector and accounting for differential aggregation of serotypes/serogroups within published studies. We aggregated serotype-specific estimates by PCV inclusion category (PCV24-additional, PCV31-additional).

**Table 1.**
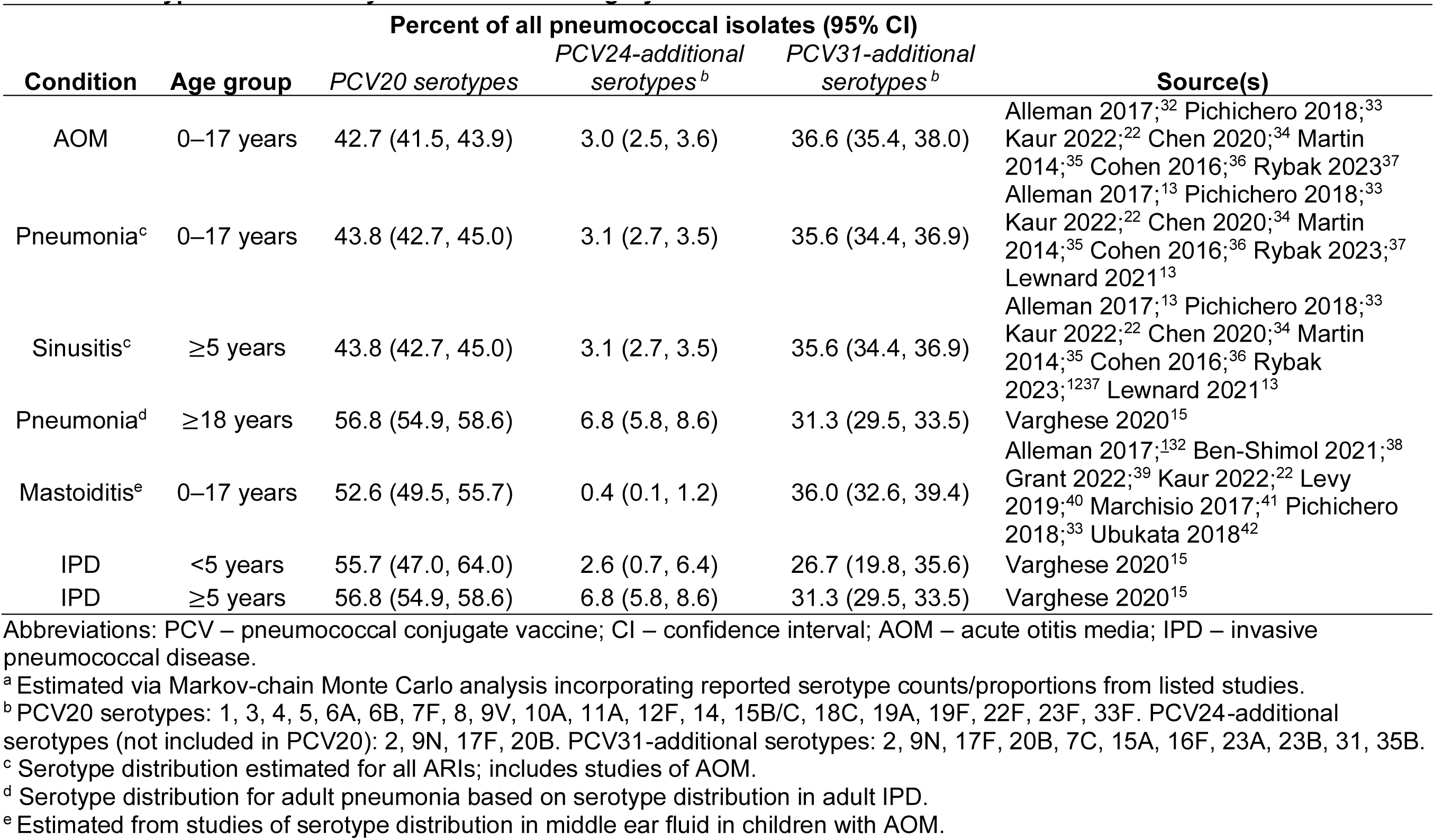
Serotype distribution by PCV inclusion category^a^.

We stratified all estimates by condition and age group (<2, 2-4, 5-17, 18-49, 50-64, and ≥65 years). Where age groups differed between our study and published estimates, we took the population-weighted average of reported estimates across age groups. For AOM, we restricted our analyses to children aged <18 years as pneumococcal AOM is primarily a childhood disease. For sinusitis, we excluded incidence in children aged <5 years, as sinusitis is infrequently diagnosed in young children and sinus development is not complete until around 5 years of age.

These analyses required simplifying assumptions given gaps in available data. First, we used adult IPD serotype distribution to extrapolate proportions of PCV24- and PCV31-additional serotypes in adult pneumococcal non-bacteremic pneumonia, as data on pneumococcal etiology in adult pneumonia cases were, with few exceptions, from studies employing urinary antigen detection of only serotypes contained in PCV13 and PCV20. Additionally, we assumed equivalent etiologic distributions for inpatient- and outpatient-managed non-bacteremic pneumonia among adults based on evidence of similar PCV13 effectiveness against all-cause pneumonia cases in both clinical settings.^13^ Last, because no studies of sinusitis etiology in adults were available post-PCV13 implementation, we assumed the same proportion of sinusitis cases were caused by pneumococci in children and adults and that serotype distribution was consistent across age groups.

### Invasive disease incidence and etiology

We obtained IPD incidence rates from the US Center for Disease Control & Prevention’s Active Bacterial Core surveillance (ABCs) from 2019.^2^ We estimated national IPD case counts by taking the product of ABCs-reported incidence and 2019 Bridged-Race census estimates^12^ by age group. We categorized IPD syndromes into mutually-exclusive categories of meningitis, bacteremic pneumonia, and other bacteremia. ABCs syndrome frequencies are not age-stratified, thus we took the following approach to distinguish proportions of each syndrome in pediatric and adult cases: we obtained pediatric proportions of IPD by syndrome from published data^14^ and used these proportions to obtain national case counts. We then subtracted pediatric case count estimates from syndrome-specific estimates of cases across all ages from 2018-19 ABCs data,^2^ as detailed in **Table S4**. Serotype distribution in IPD (stratified by <5 years and ≥5 years of age) was extracted from published ABCs data^15^ and uncertainty was propagated via Markov Chain Monte Carlo sampling, defining serotype-specific counts as a multinomial random variable for which the underlying serotype distribution was a Dirichlet parameter vector.

### Health-economic burden attributable to PCV24- and PCV31-additional serotypes

We accounted for medical and societal costs when estimating health-economic burden attributable to PCV24- and PCV31-additional serotypes. We conducted a literature review to identify published estimates for all model inputs, summarized in **Table S5**. Costs for outpatient-managed ARIs included: outpatient visit costs, stratified by emergency department (ED) and office settings; antibiotic prescription costs; patient out-of-pocket costs (e.g., non-prescription medications, transportation); surgical costs (for patients requiring surgery for complex AOM and sinusitis); missed days of work for patients or caregivers (for patients <18 years of age); and costs related to quality-adjusted life year (QALY) losses during acute illness episodes. For inpatient-managed disease (mastoiditis, non-bacteremic pneumonia, and IPD) we included: hospitalization costs; missed work during hospitalization for patients or caregivers; QALY losses during acute illness and due to premature mortality or long-term disability; and post-acute care costs. Sequelae contributing to long-term disability included deafness and neurological impairment following pneumococcal meningitis. Including societal costs associated with both QALY losses and productivity losses is consistent with recommendations for cost-effectiveness analyses.^16,17^

Across all conditions, we applied 3% annual discounting to QALY and productivity losses due to death or sequelae. We multiplied QALY losses by the 2022 US Gross Domestic Product (GDP) per capita to obtain economic valuation of QALYs; this approach aligned with the willingness-to-pay threshold endorsed as signifying “highly cost-effective” interventions by the World Health Organization CHOICE framework (Choosing Interventions that are Cost Effective).^18^ All costs were scaled using the Consumer Price Index to reflect 2022 values.

We conducted sensitivity analyses to evaluate the impact of selected model parameters and inputs on estimates, including: 1) the probability of hospitalization in IPD; 2) the probability of disability from meningitis; 3) the proportion of ARIs managed in office or ED settings; 4) the proportion of sinusitis and pediatric pneumonia attributable to pneumococcus; and 5) hospitalization costs. We present sources for estimates of each of these parameters in **Table S6**.

Data from prior published studies meet the definition of non-human subjects research according to the policies of the UC Berkeley Committee for the Protection of Human Subjects and thus Institutional Review Board review was not required.

## RESULTS

### Incidence of all-cause ARIs

Episodes of AOM, sinusitis, and non-bacteremic pneumonia caused 166.2 (95% confidence interval: 129.9-208.9), 50.3 (34.8-69.9), and 16.9 (12.0-23.0) outpatient visits per 1000 person-years among children, respectively, along with 144.1 (118.0-173.9), 46.1 (32.0-64.0), and 16.8 (13.1-21.1) antibiotic prescriptions per 1000 person-years among children (**Table S1**). Incidence of AOM and non-bacteremic pneumonia was highest among children aged <2 years.

Non-bacteremic pneumonia caused 5.4 (4.2-6.7) and 19.3 (14.1-25.6) outpatient visits per 1000 person-years among adults aged 18-64 and ≥65 years, respectively, as well as 2.9 (2.0-4.0) and 8.4 (5.4-12.2) antibiotic prescriptions per 1000 person-years at the same ages. Incidence of sinusitis was considerably higher, with 81.5 (74.6-88.9), 90.7 (82.7-99.3), and 43.4 (34.4-53.9) outpatient visits and 56.8 (51.1-63.0), 60.4 (53.9-67.4), and 24.8 (18.1-33.1) antibiotic prescriptions per 1000 person-years among adults aged 18-49, 50-64, and ≥65 years.

Incidence rates of hospital admissions for non-bacteremic pneumonia were highest among children aged <2 years (4.4 [4.2-4.6] per 1,000 person-years), adults aged 50-64 years (4.6 [4.4-4.8] per 1,000 person-years), and adults aged ≥65 years (19.1 [18.7-19.5] per 1,000 persons-years; **Table S2**). In addition, children aged <2 years experienced 0.0018 (0.0016-0.0020) hospital admissions due to mastoiditis per 1,000 person-years.

### Proportion of ARIs attributable to pneumococcus and PCV24/31-additional serotypes

Among children, pneumococcal infections accounted for 19.4% (16.8-22.2%), 27.1% (17.4-38.5%), and 45.7% (28.4-63.7%) of outpatient-managed AOM, pneumonia, and sinusitis, respectively, and 11.5% (4.6-22.3% of inpatient-managed non-bacteremic pneumonia (**Table S3**). Among adults, 9.2-10.7% of non-bacteremic pneumonia and 45.7% (28.4-63.7%) of sinusitis episodes were pneumococcal-attributable. Across all ages, these attributable fractions correspond to incidence rates of 32.3 (25.5-41.0), 32.6 (20.2-45.9), and 1.1 (0.8-1.5) outpatient-managed cases of pneumococcal AOM, sinusitis, and pneumonia, respectively, per 1000 person-years, and 0.5 (0.5-0.6) cases of hospitalized non-bacteremic pneumococcal pneumonia per 1,000 person-years (**Table S7**).

Additional (non-PCV20) serotypes in PCV24 accounted for 3.0% (2.5-3.6%) of pneumococcal AOM cases and 3.1% (2.7-3.5%) of other pneumococcal ARIs among children, as well as 0.4% (0.1-1.2%) of pneumococcal mastoiditis cases (**Table 1**). Among adults, 3.1% (2.7-3.5%) of pneumococcal sinusitis cases and 6.8% (5.8-8.6%) of pneumococcal non-bacteremic pneumonia cases were attributable to PCV24-additional serotypes. In contrast, PCV31-additional serotypes accounted for >30% of pneumococcal disease cases across non-invasive conditions and age groups.

### All-cause IPD incidence and proportion of IPD due to PCV24/31-additional serotypes

Extrapolated from 2019 ABCs age-specific data, there were an estimated 0.094 IPD cases per 1,000 person-years (**Table S8**). The highest incidence was observed in adults aged ≥65 years and 50-64 years (0.24 and 0.16 cases per 1,000 person-years) and children <2 years (0.12 cases per 1,000 person-years). Among children aged <5 years, PCV24-additional and PCV31-additional serotypes accounted for 2.6% (0.7-6.4%) and 26.7% (19.8-35.6%) of IPD cases, respectively. Across all other ages (≥5 years), PCV24-additional and PCV31-additional serotypes accounted for 6.8% (5.8-8.6%) and 31.3% (29.5-33.5%) of IPD cases.

### Health-economic burden due to PCV24-additional serotypes

Accounting for all ARI conditions, PCV24-additional serotypes caused 1.2 (0.8-1.7) outpatient visits and 0.9 (0.6-1.2) antibiotic prescriptions per 1000 person-years (**Table S9**), translating to 403,463 (277,637-550,975) outpatient visits and 293,196 (203,756-399,100) antibiotic prescriptions annually (**Table 2**). In addition, PCV24-additional serotypes caused 0.034 (0.029-0.043) hospital admissions for pneumonia and <0.001 hospitalizations for mastoiditis per 1,000 person-years or 11,145 (9,470-14,060) hospitalizations in total (**Table 2**; **Table S9**). Adults ≥65 years and 50-64 years accounted for the majority of non-bacteremic pneumonia hospitalizations with 6,461 (5,429-8,275) and 2,112 (1,767-2,709) hospitalizations, respectively.

**Table 2.**
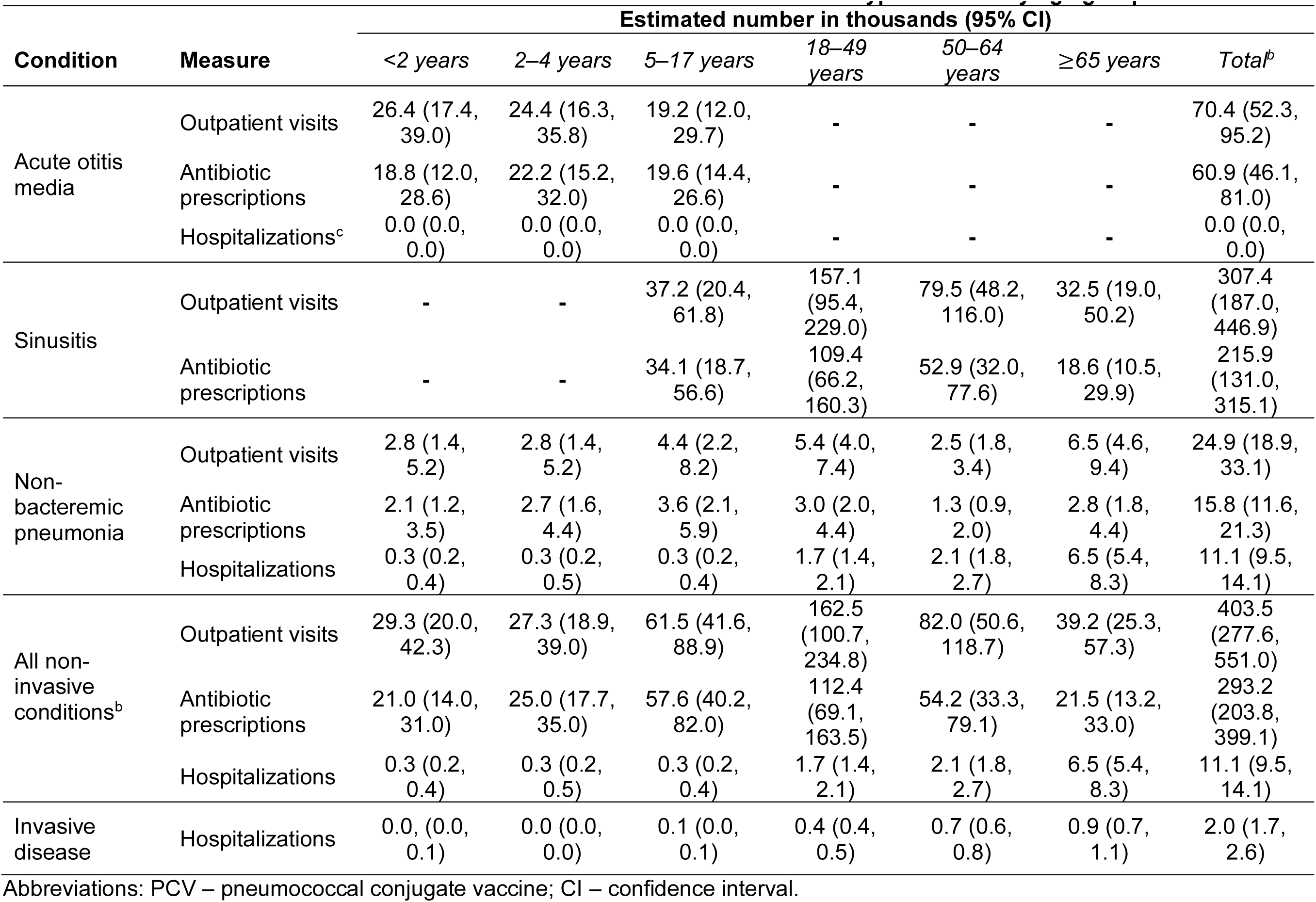
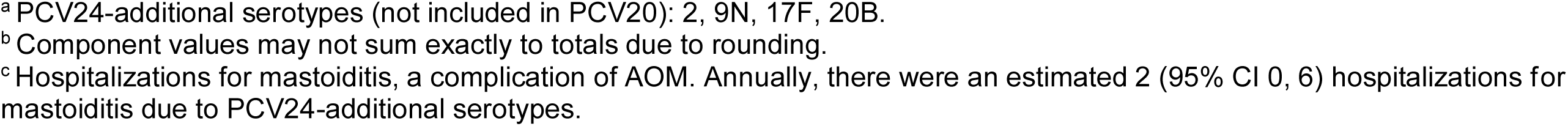
Estimated annual national burdens attributable to PCV24-additional serotype^a^ disease by age group.

| Condition | Measure | Estimated number in thousands (95% CI) |  |  |  |  |  | Total<sup>b</sup> |
| --- | --- | --- | --- | --- | --- | --- | --- | --- |
|  |  | <2 years | 2–4 years | 5–17 years | 18–49 years | 50–64 years | ≥65 years |  |
| Acute otitis media | Outpatient visits | 26.4 (17.4, 39.0) | 24.4 (16.3, 35.8) | 19.2 (12.0, 29.7) | - | - | - | 70.4 (52.3, 95.2) |
|  | Antibiotic prescriptions | 18.8 (12.0, 28.6) | 22.2 (15.2, 32.0) | 19.6 (14.4, 26.6) | - | - | - | 60.9 (46.1, 81.0) |
|  | Hospitalizations<sup>c</sup> | 0.0 (0.0, 0.0) | 0.0 (0.0, 0.0) | 0.0 (0.0, 0.0) | - | - | - | 0.0 (0.0, 0.0) |
| Sinusitis | Outpatient visits | - | - | 37.2 (20.4, 61.8) | 157.1 (95.4, 229.0) | 79.5 (48.2, 116.0) | 32.5 (19.0, 50.2) | 307.4 (187.0, 446.9) |
|  | Antibiotic prescriptions | - | - | 34.1 (18.7, 56.6) | 109.4 (66.2, 160.3) | 52.9 (32.0, 77.6) | 18.6 (10.5, 29.9) | 215.9 (131.0, 315.1) |
| Non-bacteremic pneumonia | Outpatient visits | 2.8 (1.4, 5.2) | 2.8 (1.4, 5.2) | 4.4 (2.2, 8.2) | 5.4 (4.0, 7.4) | 2.5 (1.8, 3.4) | 6.5 (4.6, 9.4) | 24.9 (18.9, 33.1) |
|  | Antibiotic prescriptions | 2.1 (1.2, 3.5) | 2.7 (1.6, 4.4) | 3.6 (2.1, 5.9) | 3.0 (2.0, 4.4) | 1.3 (0.9, 2.0) | 2.8 (1.8, 4.4) | 15.8 (11.6, 21.3) |
|  | Hospitalizations | 0.3 (0.2, 0.4) | 0.3 (0.2, 0.5) | 0.3 (0.2, 0.4) | 1.7 (1.4, 2.1) | 2.1 (1.8, 2.7) | 6.5 (5.4, 8.3) | 11.1 (9.5, 14.1) |
| All non-invasive conditions<sup>b</sup> | Outpatient visits | 29.3 (20.0, 42.3) | 27.3 (18.9, 39.0) | 61.5 (41.6, 88.9) | 162.5 (100.7, 234.8) | 82.0 (50.6, 118.7) | 39.2 (25.3, 57.3) | 403.5 (277.6, 551.0) |
|  | Antibiotic prescriptions | 21.0 (14.0, 31.0) | 25.0 (17.7, 35.0) | 57.6 (40.2, 82.0) | 112.4 (69.1, 163.5) | 54.2 (33.3, 79.1) | 21.5 (13.2, 33.0) | 293.2 (203.8, 399.1) |
|  | Hospitalizations | 0.3 (0.2, 0.4) | 0.3 (0.2, 0.5) | 0.3 (0.2, 0.4) | 1.7 (1.4, 2.1) | 2.1 (1.8, 2.7) | 6.5 (5.4, 8.3) | 11.1 (9.5, 14.1) |
| Invasive disease | Hospitalizations | 0.0, (0.0, 0.1) | 0.0 (0.0, 0.0) | 0.1 (0.0, 0.1) | 0.4 (0.4, 0.5) | 0.7 (0.6, 0.8) | 0.9 (0.7, 1.1) | 2.0 (1.7, 2.6) |

Abbreviations: PCV – pneumococcal conjugate vaccine; CI – confidence interval.
<sup>a</sup> PCV24-additional serotypes (not included in PCV20): 2, 9N, 17F, 20B.
<sup>b</sup> Component values may not sum exactly to totals due to rounding.
<sup>c</sup> Hospitalizations for mastoiditis, a complication of AOM. Annually, there were an estimated 2 (95% CI 0, 6) hospitalizations for mastoiditis due to PCV24-additional serotypes.

Incidence of IPD due to PCV24-additional serotypes was 0.006 (0.005-0.008) per 1,000 person-years (**Table S8**), equivalent to 2,035 (1,743-2,587) IPD cases annually (**Table 2**). Of this total, 868 (745-1104) and 666 (572-848) IPD cases due to PCV24-additional serotype were expected to have occurred among adults aged ≥65 years and 50-64 years, respectively (**Table 2**).

Total annual health-economic burden attributable to PCV24-additional serotype infections amounted to $1.3 billion ($1.1-1.7b) in 2022 USD (**Table 3**). Inpatient-attended disease (invasive and non-invasive) accounted for $1.1b ($0.9-1.4b) of this total cost. Non-bacteremic pneumonia was the largest driver of inpatient costs at $690.5 million ($566.5-$902.4m), most of which came from adults aged ≥65 years ($475.8m [$381.0-$641.3m]) and 50-64 years ($124.7m [$97.3-$173.8m]; **Table S10**). Non-bacteremic pneumonia also resulted in 6,863 (5,817-8,756) years of life lost due to premature death (**Table S11**), resulting is costs of $524.3m ($444.4-668.9m) (**Table S10**). Costs from IPD due to PCV24-additional serotypes totaled $411.3m ($348.1-525.9m), including $289.9m ($248.5-368.6m) resulting from 3,794 (3,253-4,825) years of life lost due to premature death (**Tables S11-S12**) and $26.6m ($22.8-33.9m) resulting from 349 (299-443) QALYs lost due to sequelae (**Tables S12**-**S13**). Outpatient-managed ARIs due to PCV24-additional serotypes accounted for $210.9m ($155.3-287.9m) in costs, with medical and non-medical costs totaling $85.3m ($52.0-138.6m) and $124.8m ($96.6-160.5m), respectively (**Tables 3**; **Table S14**). Of the total costs for outpatient-managed ARIs due to PCV24-additional serotypes, infections among children aged <2 years cost $29.2m ($19.6-43.1m) and those in adults aged ≥65 years totaled $27.7m ($20.0-38.6m).

**Table 3.**
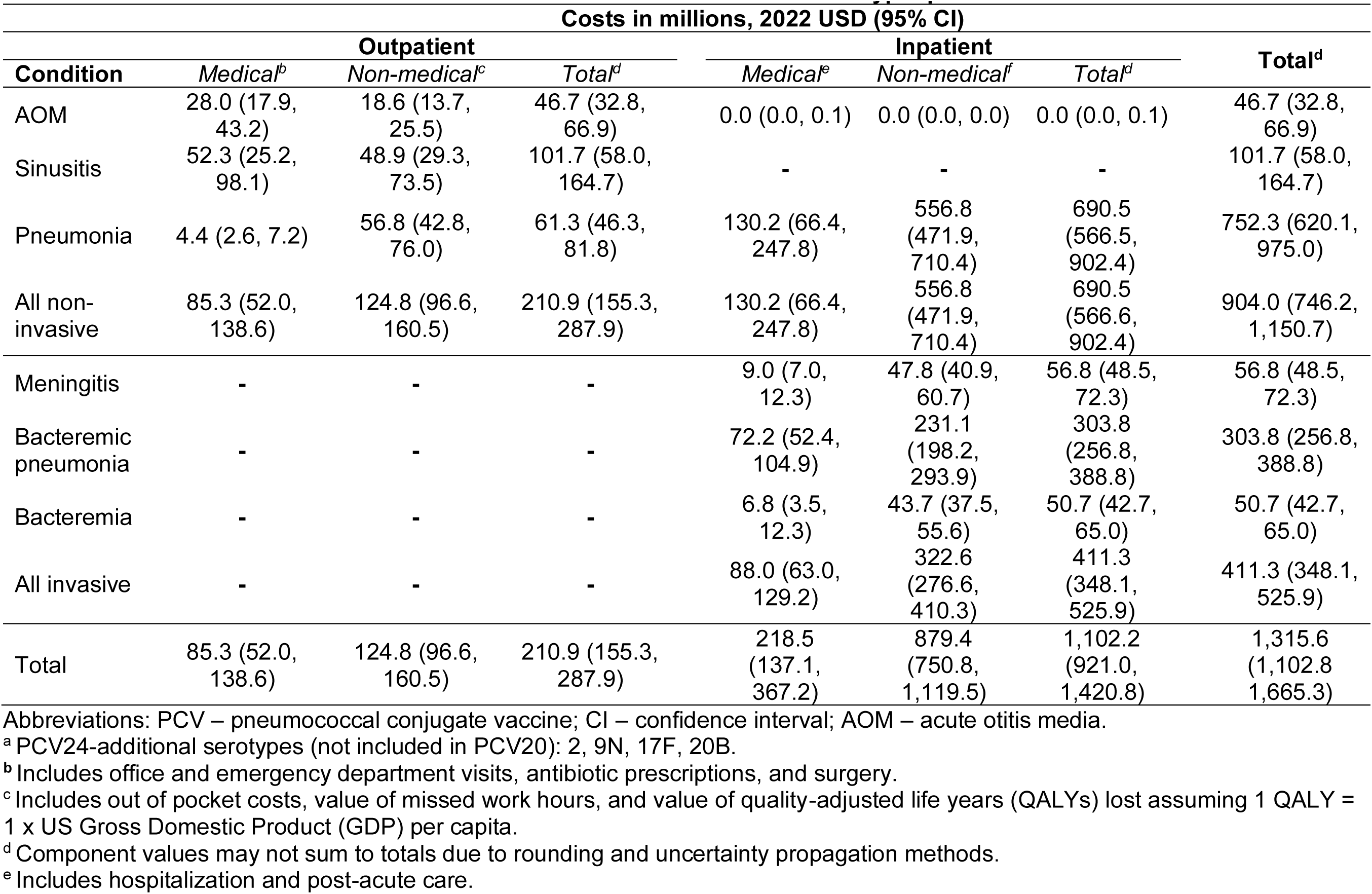
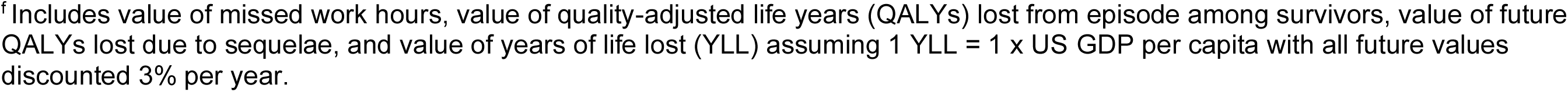
Annual health economic burden attributable to PCV24-additional serotype^a^ pneumococcal disease.

| Costs in millions, 2022 USD (95% CI) |  |  |  |  |  |  |  |
| --- | --- | --- | --- | --- | --- | --- | --- |
| Condition | Outpatient |  |  | Inpatient |  |  | Total<sup>d</sup> |
|  | Medical<sup>b</sup> | Non-medical<sup>c</sup> | Total<sup>d</sup> | Medical<sup>e</sup> | Non-medical<sup>f</sup> | Total<sup>d</sup> |  |
| AOM | 28.0 (17.9, 43.2) | 18.6 (13.7, 25.5) | 46.7 (32.8, 66.9) | 0.0 (0.0, 0.1) | 0.0 (0.0, 0.0) | 0.0 (0.0, 0.1) | 46.7 (32.8, 66.9) |
| Sinusitis | 52.3 (25.2, 98.1) | 48.9 (29.3, 73.5) | 101.7 (58.0, 164.7) | - | - | - | 101.7 (58.0, 164.7) |
| Pneumonia | 4.4 (2.6, 7.2) | 56.8 (42.8, 76.0) | 61.3 (46.3, 81.8) | 130.2 (66.4, 247.8) | 556.8 (471.9, 710.4) | 690.5 (566.5, 902.4) | 752.3 (620.1, 975.0) |
| All non-invasive | 85.3 (52.0, 138.6) | 124.8 (96.6, 160.5) | 210.9 (155.3, 287.9) | 130.2 (66.4, 247.8) | 556.8 (471.9, 710.4) | 690.5 (566.6, 902.4) | 904.0 (746.2, 1,150.7) |
| Meningitis | - | - | - | 9.0 (7.0, 12.3) | 47.8 (40.9, 60.7) | 56.8 (48.5, 72.3) | 56.8 (48.5, 72.3) |
| Bacteremic pneumonia | - | - | - | 72.2 (52.4, 104.9) | 231.1 (198.2, 293.9) | 303.8 (256.8, 388.8) | 303.8 (256.8, 388.8) |
| Bacteremia | - | - | - | 6.8 (3.5, 12.3) | 43.7 (37.5, 55.6) | 50.7 (42.7, 65.0) | 50.7 (42.7, 65.0) |
| All invasive | - | - | - | 88.0 (63.0, 129.2) | 322.6 (276.6, 410.3) | 411.3 (348.1, 525.9) | 411.3 (348.1, 525.9) |
| Total | 85.3 (52.0, 138.6) | 124.8 (96.6, 160.5) | 210.9 (155.3, 287.9) | 218.5 (137.1, 367.2) | 879.4 (750.8, 1,119.5) | 1,102.2 (921.0, 1,420.8) | 1,315.6 (1,102.8, 1,665.3) |

Abbreviations: PCV – pneumococcal conjugate vaccine; CI – confidence interval; AOM – acute otitis media.
<sup>a</sup> PCV24-additional serotypes (not included in PCV20): 2, 9N, 17F, 20B.
<sup>b</sup> Includes office and emergency department visits, antibiotic prescriptions, and surgery.
<sup>c</sup> Includes out of pocket costs, value of missed work hours, and value of quality-adjusted life years (QALYs) lost assuming 1 QALY = 1 x US Gross Domestic Product (GDP) per capita.
<sup>d</sup> Component values may not sum to totals due to rounding and uncertainty propagation methods.
<sup>e</sup> Includes hospitalization and post-acute care.
<sup>f</sup> Includes value of missed work hours, value of quality-adjusted life years (QALYs) lost from episode among survivors, value of future QALYs lost due to sequelae, and value of years of life lost (YLL) assuming 1 YLL = 1 x US GDP per capita with all future values discounted 3% per year.

### Health-economic burden due to PCV31-additional serotypes

In total, ARIs attributable to PCV31-additional serotypes resulted in 14.2 (9.9-18.7) outpatient visits and 10.4 (7.4-13.6) antibiotic prescriptions per 1000 person-years (**Table S9**), equivalent to 4,651,292 (3,259,355-6,147,664) visits and 3,409,232 (2,424,662-4,479,297) antibiotic prescriptions annually (**Table 4**). Additionally, 57,750 (52,154-64,015) and 165 (91-256) hospitalizations for non-bacteremic pneumonia and mastoiditis, respectively, were due to PCV31-additional serotype-attributable disease, corresponding to 17.6 (15.9-19.5) and 0.2 (0.1-0.3) cases per 100,000 person-years (**Table S9**). Among adults ≥65 years, there were an estimated 29,816 (26,941-33,043) hospitalizations for non-bacteremic pneumonia (**Table 4**).

**Table 4.** Estimated annual national burdens attributable to PCV31-additional serotype^a^ disease by age group.

| Condition | Measure | Estimated number in thousands (95% CI) |  |  |  |  |  | Total<sup>b</sup> |
| --- | --- | --- | --- | --- | --- | --- | --- | --- |
|  |  | <2 years | 2–4 years | 5–17 years | 18–49 years | 50–64 years | ≥65 years |  |
| Acute otitis media | Outpatient visits | 323.9<br>(220.9, 459.4) | 299.5<br>(207.1, 420.3) | 235.9<br>(151.5, 350.7) | - | - | - | 865.7<br>(672.4, 1,101.3) |
|  | Antibiotic prescriptions | 231.7<br>(152.3, 338.1) | 272.9<br>(193.7, 374.8) | 240.4<br>(184.7, 308.9) | - | - | - | 748.9<br>(596.0, 933.6) |
|  | Hospitalizations<sup>c</sup> | 0.0 (0.0, 0.1) | 0.0 (0.0, 0.0) | 0.1 (0.1, 0.2) | - | - | - | 0.2 (0.1, 0.3) |
| Sinusitis | Outpatient visits | - | - | 433.9<br>(241.6, 706.1) | 1,834.8<br>(1,132.3, 2,597.2) | 928.5<br>(572.4, 1,315.9) | 379.6<br>(225.7, 571.9) | 3,590.9<br>(2,218.7, 5,066.0) |
|  | Antibiotic prescriptions | - | - | 397.8<br>(221.7, 646.5) | 1,278.0<br>(786.1, 1,819.0) | 617.4<br>(379.4, 880.7) | 216.6<br>(124.0, 341.8) | 2,522.1<br>(1,555.5, 3,574.0) |
| Non-bacteremic pneumonia | Outpatient visits | 33.0 (16.7, 59.9) | 32.9 (16.9, 59.3) | 51.7 (26.2, 93.7) | 24.9 (19.2, 31.8) | 11.3 (8.7, 14.5) | 30.1 (21.7, 40.5) | 186.7<br>(135.3, 258.0) |
|  | Antibiotic prescriptions | 24.9 (14.2, 40.5) | 32.0 (19.1, 50.1) | 41.8 (24.4, 67.1) | 13.6 (9.4, 19.0) | 6.2 (4.3, 8.6) | 13.0 (8.4, 19.3) | 132.9<br>(93.8, 182.7) |
|  | Hospitalizations | 3.2 (2.1, 4.6) | 3.6 (2.3, 5.2) | 3.5 (2.3, 5.1) | 7.7 (6.9, 8.6) | 9.7 (8.7, 10.9) | 29.8 (26.9, 33.0) | 57.8 (52.2, 64.0) |
| All non-invasive conditions<sup>b</sup> | Outpatient visits | 358.4<br>(253.0, 495.7) | 333.9<br>(238.8, 456.6) | 728.7<br>(508.1, 1,020.9) | 1859.8<br>(1,157.2, 2,622.5) | 939.9<br>(583.8, 1,327.3) | 410.2<br>(255.6, 602.6) | 4,651.3<br>(3,259.4, 6,147.7) |
|  | Antibiotic prescriptions | 257.2<br>(176.7, 364.5) | 305.7<br>(224.7, 408.8) | 683.3<br>(493.8, 940.5) | 1,291.7<br>(799.7, 1,832.8) | 623.6<br>(385.7, 887.0) | 229.9<br>(137.1, 355.2) | 3,409.2<br>(2,424.7, 4,479.3) |
|  | Hospitalizations | 3.2 (2.1, 4.6) | 3.6 (2.3, 5.2) | 3.7 (2.4, 5.2) | 7.7 (6.9, 8.6) | 9.7 (8.7, 10.9) | 29.8 (26.9, 33.0) | 57.9 (52.3, 64.2) |
|  |  | <2 years | 2–4 years | 5–17 years | 18–49 years | 50–64 years | ≥65 years |  |
| Invasive disease | Hospitalizations | 0.2 (0.2, 0.3) | 0.1 (0.1, 0.2) | 0.2 (0.2, 0.3) | 1.9 (1.8, 2.0) | 3.1 (2.9, 3.3) | 4.0 (3.8, 4.3) | 9.6 (9.0, 10.3) |

Abbreviations: PCV – pneumococcal conjugate vaccine; CI – confidence interval; AOM – acute otitis media.
<sup>a</sup> PCV31-additional serotypes: 2, 9N, 17F, 20B, 7C, 15A, 16F, 23A, 23B, 31, 35B.
<sup>b</sup> Component values may not sum exactly to totals due to rounding.
<sup>c</sup> Hospitalizations for mastoiditis, a complication of AOM.

Incidence of IPD due to PCV31-additional serotypes was 0.029 (0.027-0.031) cases per 1,000 person-years, (**Table S8**), equivalent to 9,593 (9,003-10,278) IPD cases annually (**Table 4**). There were an estimated 4,006 (3,772-4,285) and 3,075 (2,896-3,290) IPD hospitalizations due to PCV31-additional serotype disease among adults ≥65 years and 50-64 years, respectively. In contrast, children <2 years accounted for 245 (181-326) cases and children 2-4 years accounted for 138 (102-183) cases.

Total annual health-economic burden attributable to PCV31-additional serotype disease was approximately $7.5b ($6.6-$8.6b) 2022 USD, with outpatient-managed ARIs contributing $2.2b ($1.7-3.0b) and inpatient-managed disease contributing $5.2b ($4.7-5.9b) in costs (**Table 5**). Non-bacteremic pneumonia alone accounted for almost half of all costs ($3.8b [$3.3-4.4b]), with adults ≥65 years responsible for $2.2b ($1.9-2.7b) and $90.1m ($65.4-121.2m) in costs from non-bacteremic pneumonia managed in inpatient and outpatient settings, respectively (**Table S15**). Non-bacteremic pneumonia due to PCV31-additional serotypes also resulted in 32,055 (29,40-35,054) years of life lost to premature death (**Table S11**), which accounted for $2.4b ($2.3-2.7b) in losses (**Table S15).** PCV31-additional serotype IPD resulted in costs of $1.9b ($1.8-2.1b) (**Table S16**). Non-medical costs accounted for $1.5b ($1.4-1.6b) of these costs, with $1.4b ($1.3-1.5b) resulting from 17,797 (16,719-19,058) years of life lost due to premature deaths (**Tables S16, S11**). Outpatient costs attributable to PCV31-additional serotypes were $2.2b ($1.7-3.0b), with medical costs of $990.6m ($615.1-$1578.5m) (**Table 5**; **Table S17**). Costs associated with outpatient-managed ARIs attributable to PCV31-additional serotypes among children aged <2 years and adults aged ≥65 years totaled $353.6 ($244.6-505.2) million and $182.9 ($129.0-262.7) million, respectively (**Table S17**).

**Table 5.** Annual health economic burden attributable to PCV31-additional serotype^a^ pneumococcal disease.

| Costs in millions, 2022 USD (95% CI) |  |  |  |  |  |  |  |
| --- | --- | --- | --- | --- | --- | --- | --- |
| Condition | Outpatient |  |  | Inpatient |  |  | Total<sup>d</sup> |
|  | Medical<sup>b</sup> | Non-medical<sup>c</sup> | Total<sup>d</sup> | Medical<sup>e</sup> | Non-medical<sup>f</sup> | Total<sup>d</sup> |  |
| AOM | 343.5 (226.8, 511.7) | 228.4 (175.4, 295.9) | 573.1 (418.8, 783.8) | 2.0 (0.5, 5.7) | 0.1 (0.1, 0.2) | 2.2 (0.6, 5.9) | 575.1 (421.1, 786.3) |
| Sinusitis | 609.7 (298.1, 1,125.5) | 570.6 (347.5, 835.3) | 1,187.2 (686.6, 1,881.2) | - | - | - | 1,187.2 (686.6, 1,881.2) |
| Pneumonia | 30.8 (18.3, 50.2) | 435.3 (311.7, 607.1) | 466.7 (335.8, 647.9) | 668.9 (376.8, 1,179.3) | 2,601.7 (2,386.6, 2,845.1) | 3,278.8 (2,878.6, 3,861.9) | 3,754.3 (3,293.8, 4,382.1) |
| All non-invasive | 990.6 (615.1, 1,578.5) | 1,241.0 (965.4, 1,568.7) | 2,239.6 (1,656.7, 3,026.0) | 671.2 (379.0, 1,181.6) | 2,601.8 (2,386.7, 2,845.2) | 3,281.3 (2,880.9, 3,864.5) | 5,538.1 (4,741.7, 6,590.5) |
| Meningitis | - | - | - | 43.2 (35.7, 54.5) | 224.8 (211.1, 240.8) | 268.4 (250.1, 290.1) | 268.4 (250.1, 290.1) |
| Bacteremic pneumonia | - | - | - | 334.3 (253.6, 458.6) | 1,083.5 (1,018.0, 1,160.2) | 1,420.4 (1,301.5, 1,575.3) | 1,420.4 (1,301.5, 1,575.3) |
| Bacteremia | - | - | - | 31.8 (16.9, 55.0) | 205.0 (192.6, 219.5) | 237.4 (216.2, 265.4) | 237.4 (216.2, 265.4) |
| All invasive | - | - | - | 409.3 (306.4, 567.9) | 1,513.3 (1,421.7, 1,620.5) | 1,926.3 (1,768.8, 2,129.2) | 1,926.3 (1,768.8, 2,129.2) |
| Total | 990.6 (615.1, 1,578.5) | 1,241.0 (965.4, 1,568.7) | 2,239.6 (1,656.7, 3,026.0) | 1,081.4 (721.7, 1,708.6) | 4,115.3 (3,829.5, 4,445.3) | 5,209.2 (4,693.3, 5,940.8) | 7,467.3 (6,586.1, 8,627.0) |

Abbreviations: PCV – pneumococcal conjugate vaccine; CI – confidence interval; AOM – acute otitis media.
<sup>a</sup> PCV31-additional serotypes: 2, 9N, 17F, 20B, 7C, 15A, 16F, 23A, 23B, 31, 35B.
<sup>b</sup> Includes office and emergency department visits, antibiotic prescriptions, and surgery.
<sup>c</sup> Includes out of pocket costs, value of missed work hours, and value of quality-adjusted life years (QALYs) lost assuming 1 QALY = 1 x US Gross Domestic Product (GDP) per capita.
<sup>d</sup> Component values may not sum to totals due to rounding and uncertainty propagation methods.
<sup>e</sup> Includes hospitalization and post-acute care.
<sup>f</sup> Includes value of missed work hours, value of quality-adjusted life years (QALYs) lost from episode among survivors, value of future QALYs lost due to sequelae, and value of years of life lost (YLL) assuming 1 YLL = 1 x US GDP per capita with all future values discounted 3% per year.

### Sensitivity analyses

Of all parameters explored, the proportion of sinusitis episodes attributed to pneumococcus had the greatest impact on health-economic burden estimates. Reducing this proportion from 45.7% to 11.8% yielded a decrease in burden of $77.9m ($31.1-138.5m) for PCV24-additional serotypes and $0.9b ($0.4-$1.6b) for PCV31-additional serotypes (**Table S6**). Sizeable changes were also observed in analyses varying hospitalization costs in non-bacteremic pneumonia; sensitivity analysis estimates for health-economic burden of PCV24-additional serotypes ranged from $72.8m below to $116.4m above our primary estimate, while for PCV31-additional serotypes, estimates ranged from $389.1m below to $716.4m above the primary estimate.

## DISCUSSION

We estimated that PCV24-additional serotypes (2, 9N, 17F, 20B) are collectively responsible for $1.3 billion 2022 USD in health-economic burden annually. Of this total, ARIs (AOM, sinusitis, non-bacteremic pneumonia) account for $904 million and IPD accounts for $411m. Novel serotypes included in PCV31 (2, 9N, 17F, 20B, 7C, 15A, 16F, 23A, 23B, 31, 35B) account for $7.5b in health-economic burden annually, with $5.5b owing to ARIs and $1.9b from IPD. The largest driver of costs across PCV inclusion categories was non-bacteremic pneumonia, particularly in adults ≥65 years and 50-64 years. These age groups also accounted for the largest share of costs relating to IPD.

The estimated health-economic burden of PCV31-additional serotypes exceeds that of PCV24-additional serotypes by almost five fold. Notably, the differential between the two PCV inclusion categories is greater in pediatric ARIs compared with adult pneumonia and invasive disease among both children and adults. A large differential in burden between the two PCV inclusion categories is expected as in pediatric ARIs, PCV31-additional serotypes account for approximately 36% of pneumococcal infections compared with 3% of pneumococcal infections caused by PCV24-additional serotypes while in adult IPD and non-bacteremic pneumonia PCV24-additional serotypes account for almost 7% of pneumococcal infections while PCV31 serotypes account for 31%. This difference may be because both PCV24 and PCV31 target serotypes 9N and 20 which contribute to IPD^15^ but are less prominent in pediatric ARIs.^1^ Inclusion of serotype 35B in PCV31 is an important contributing factor in the difference between PCV24 and PCV31, given the growing importance of 35B in IPD,^19^ carriage in children,^20^ and AOM.^21,22^ Longstanding surveillance of otopathogens in middle ear fluid^21^ and IPD^23^ identified increases in 35B virulence associated with lineage shifts in the post-PCV13 period. Given the additional problem of antibiotic nonsusceptibility in 35B isolates,^19,24,25^ this serotype is an important target for vaccines as well as ongoing surveillance. In addition to 35B, 23A is unique to PCV31 and is an important contributor to IPD,^15^ adult pneumonia,^26^ and pediatric ARIs.^1^

Although preventing IPD is a major focus of pneumococcal vaccine development and informs vaccine recommendations,^10^ our findings demonstrate large potential health-economic benefits from preventing non-bacteremic pneumonia, especially in adults aged ≥50 years. However, data on serotype distribution in this clinical condition is limited. The most expansive serotype-specific urinary antigen detection (SSUAD) tests are restricted to 24^27^ and 31^26^ serotypes and do not include all PCV31-additional serotypes and other non-vaccine type serotypes. Development of SSUAD tests capturing a wide range of serotypes, along with studies of differences in UAD sensitivity by serotype and pneumococcal vaccination status, remain needed to inform understanding of serotype distribution in non-bacteremic pneumonia.

Current guidelines recommend that all PCV-naïve adults aged ≥65 years and adults aged <65 with certain underlying risk factors receive PCV15 and PPSV23 or PCV20.^10^ Adults aged 50-64 years without underlying medical conditions are not recommended to receive PCVs,^10^ although including this age group in future PCV recommendations is under consideration.^28^ Our study demonstrated the highest non-bacteremic pneumonia and IPD burdens in adults aged ≥65 years and 50-64 years, suggesting preventing pneumococcal disease in the 50-64 year age group may be associated with appreciable health-economic impact.

In contrast, IPD due to PCV24/31-additional serotypes was rare among young children, who are recommended to receive PCVs. The greatest benefits of PCV24 and PCV31 in children may come through preventing ARIs. Children <2 years experienced the second-highest incidence of pneumonia-related hospitalization (following adults ≥65 years) and the highest incidence of pneumonia-related outpatient visits. Additionally, incidence rates of ARI-related outpatient visits and antibiotic prescriptions were highest among children aged <2 years, followed by children aged 2-4 years. Within these age groups, PCV24- and PCV31-additional serotypes accounted for 46,000 and 563,000 antibiotic prescriptions annually, respectively, representing 0.3% and 3.2% of all antibiotic use, assuming a total of 17.9m antibiotic prescriptions dispensed to children annually in the United States.^1^

Our study has limitations. First, we relied on published estimates for all inputs; thus, our study is restricted by available data and subject to limitations of included studies. Literature gaps concerning etiology and pneumococcal serotype distributions are greatest for sinusitis and non-bacteremic pneumonia. We used data from other conditions to inform our estimates and include sensitivity analyses to evaluate the impact of varying etiologic assumptions. Second, our study describes only the burden associated with PCV24/31-additional serotypes; as clinical effectiveness of these vaccines remains to be determined,^29^ we do not estimate vaccine-preventable burden or cost-effectiveness of PCV24/31 programs. Third, our valuation of QALY losses assumes a single willingness-to-pay threshold (GDP per capita). In practice, this threshold is not universal,^30^ and in the United States, some recommended vaccinations exceed WHO cost-effectiveness thresholds (e.g., serogroup B meningococcal vaccine^31^). Fourth, our estimates are based on non-PCV20 serotypes targeted by PCV24/31 and we did not compare burdens from PCV24/31 and PCV15 serotypes, although differences would be expected to be even greater than those observed in our study. Last, our study relied on data from 2019 and earlier to capture disease burden and healthcare utilization during periods not subject to major disruption from the COVID-19 pandemic. It remains to be determined if pneumococcal disease burden will align with pre-pandemic observations in the current context of endemic SARS-CoV-2 circulation.

Non-PCV20 serotypes in PCV24 and PCV31, especially those in PCV31, impose substantial health-economic burden in the United States. If determined to be safe and immunogenic in ongoing trials, these vaccines may contribute to substantial reductions in pneumococcal disease, associated morbidity and mortality, and economic costs. Monitoring serotype-specific residual incidence of pneumococcal ARIs and IPD, and resulting health-economic burden, remains necessary to optimize PCV formulations and recommendations for differing ages and risk groups.

## Data Availability

All data produced in the present work are contained in the manuscript.

