## Supplemental Materials for "Health-economic burden attributable to novel serotypes in candidate 24- and 31-valent pneumococcal conjugate vaccines"

**Table S1.** All-cause outpatient incidence estimates

**Table S2.** All-cause non-invasive hospitalization incidence estimates

**Table S3.** Proportion of non-invasive cases attributable to *Streptococcus pneumoniae*

**Table S4.** Estimated proportions of IPD cases by syndrome from 2018–2019 ABCs data and Prasad et al. 2023

**Table S5.** Health economic burden model inputs

**Table S6.** Sensitivity analyses

**Table S7.** Estimated incidence and national annual burdens attributable to non-invasive pneumococcal infections (all serotypes)

**Table S8.** Estimated incidence of invasive pneumococcal disease attributable to PCV24- and PCV31-additional serotypes, 2019

**Table S9.** Estimated incidence of healthcare utilization attributable to non-invasive PCV24- and PCV31-additional serotype disease

**Table S10.** Annual inpatient health economic burden attributable to non-invasive PCV24-additional serotype pneumococcal disease

**Table S11.** Years life lost due to pneumococcal disease from PCV24- and PCV31-additional serotypes

**Table S12.** Annual health economic burden attributable to invasive PCV24-additional serotype pneumococcal disease

**Table S13.** Quality adjusted life years lost due to sequelae of pneumococcal meningitis from PCV24- and PCV31-additional serotypes

**Table S14.** Annual outpatient health economic burden attributable to non-invasive PCV24-additional serotype pneumococcal disease

**Table S15.** Annual inpatient health economic burden attributable to non-invasive PCV31-additional serotype pneumococcal disease

**Table S16.** Annual health economic burden attributable to invasive PCV31-additional serotype pneumococcal disease

**Table S17.** Annual outpatient health economic burden attributable to non-invasive PCV31-additional serotype pneumococcal disease

**Supplemental Material References**

**Table S1. All-cause outpatient incidence estimates**

| Condition | Age group | Incidence per 1000 person-years (95% CI)[a] | | Source |
|---|---|---|---|---|
| | | *Outpatient visits* | *Antibiotic prescriptions* | |
| AOM | <2 years | 601.0 (421.0, 825.8) | 429.8 (289.8, 609.3) | King et al. 2024[1] |
| | 2–4 years | 353.2 (251.3, 479.6) | 321.8 (235.7, 426.7) | King et al. 2024[1] |
| | 5–17 years | 62.2 (40.9, 89.9) | 63.4 (50.8, 77.8) | King et al. 2024[1] |
| Pneumonia | <2 years | 45.5 (26.6, 71.8) | 34.1 (23.9, 46.9) | King et al. 2024[1] |
| | 2–4 years | 28.8 (17.1, 45.1) | 27.9 (21.1, 36.0) | King et al. 2024[1] |
| | 5–17 years | 10.1 (5.9, 16.0) | 8.2 (5.9, 10.9) | King et al. 2024[1] |
| | 18–49 years[b] | 5.4 (4.2–6.7) | 2.9 (2.0, 4.0) | Hersh et al. 2021[2] |
| | 50–64 years[b] | 5.4 (4.2–6.7) | 2.9 (2.0, 4.0) | Hersh et al. 2021[2] |
| | ≥65 years | 19.3 (14.1–25.6) | 8.4 (5.4, 12.2) | Hersh et al. 2021[2] |
| Sinusitis | 5–17 years | 50.3 (34.8, 69.9) | 46.1 (32.0, 64.0) | King et al. 2024[1] |
| | 18–49 years[c] | 81.5 (74.6, 88.9) | 56.8 (51.1, 63.0) | Lewnard et al. 2021[3] |
| | 50–64 years[d] | 90.7 (82.7, 99.3) | 60.4 (53.9, 67.4) | Lewnard et al. 2021[3] |
| | ≥65 years | 43.4 (34.4, 53.9) | 24.8 (18.1, 33.1) | Hersh et al. 2021[2] |

Abbreviations: CI – confidence interval; AOM – acute otitis media.
[a] Incidence estimates presented obtained by fitting a gamma distribution to reported estimates (point estimate and uncertainty intervals) from published studies and estimating the median and 95% confidence limits from five million draws from the fitted distribution.
[b] Original study age group was 20–64 years; extrapolated to ages 18-19 years for analysis. Hersh et al. 2021[2] reported a single incidence estimate for all individuals 18–64 years, which we applied to both of the corresponding age groups (18–49 years and 50–64 years) in our study.
[c] Obtained by estimating the population-weighted average of reported incidence values for 18–39 years and 40–64 years from Lewnard et al. 2021.[3]
[d] Lewnard et al. 2021[3] report incidence for individuals 40–64 years. We apply that estimate to analyses for our study age group of 50–64 years.

**Table S2. All-cause non-invasive hospitalization incidence estimates**

| Condition | Age group | Hospitalizations per 100,000 person-years (95% CI)[a] | Source |
|---|---|---|---|
| Mastoiditis | <2 years | 3.4 (2.9, 4.0) | King et al. 2019[4] |
| | 2–4 years[b] | 1.6 (1.4, 1.8) | King et al. 2019[4] |
| | 5–17 years[b] | 1.6 (1.4, 1.8) | King et al. 2019[4] |
| Pneumonia | <2 years[c] | 439.3 (423.8, 455.2) | Hayes et al. 2018[5] |
| | 2–4 years[d] | 314.3 (300.1, 329.0) | Hayes et al. 2018[5] |
| | 5–17 years[e] | 68.7 (65.1, 72.5) | Hayes et al. 2018[5] |
| | 18–49 years[f] | 166.2 (160.4, 172.1) | Hayes et al. 2018[5] |
| | 50–64 years[g] | 461.9 (440.9, 483.7) | Hayes et al. 2018[5] |
| | ≥65 years[h] | 1913.6 (1,873.5, 1,954.4) | Hayes et al. 2018[5] |

Abbreviations: CI – confidence interval.
[a] Incidence estimates presented obtained by fitting gamma distributions to reported estimates (point estimate and uncertainty intervals) from published studies and estimating the median and 95% confidence limits from five million draws from the fitted distribution.
[b] King et al. 2019[4] reported a single incidence estimate for all individuals 2–17 years, which we applied to both of the corresponding age groups (2–4 years and 5–17 years) in our study.
[c] Obtained by estimating the population-weighted average of reported incidence values for children aged <1 year and 1–4 years (for children aged 1 year) from Hayes et al. 2018.[5]
[d] Estimated based on reported incidence value for children 1–4 years from Hayes et al. 2018.[5]
[e] Estimated based on reported incidence value for children 5–19 years from Hayes et al. 2018.[5]
[f] Obtained by estimating the population-weighted average of reported incidence values for age groups 5–19 years, 20–44 years, and 45–64 years from Hayes et al. 2018.[5]
[g] Estimated based on reported incidence value for adults 45–64 years from Hayes et al. 2018.[5]
[h] Obtained by estimating the population-weighted average of reported incidence values for age groups 65–74 years, 75–84 years, and ≥85 years from Hayes et al. 2018.[5]

**Table S3. Proportion of non-invasive cases attributable to *Streptococcus pneumoniae***

| Setting | Condition | Age group | Pneumococcal-attributable percent (95% CI)[a] | Source |
|---|---|---|---|---|
| Outpatient | AOM | 0–17 years | 19.4 (16.8–22.2) | King et al. 202[1] |
| | Pneumonia | 0–17 years | 27.1 (17.4–38.5) | King et al. 2024[1] |
| | | 18–64 years | 10.7 (9.9–11.5)[b] | Isturiz et al. 2019[6] |
| | | ≥65 years | 9.2 (8.5–9.9)[b] | Isturiz et al. 2019[6] |
| | Sinusitis | 5–17 years | 45.7 (28.4–63.7) | King et al. 2024[1] |
| | | ≥18 years | 45.7 (28.4–63.7)[c] | King et al. 2024[1] |
| Inpatient | Mastoiditis | 0–17 years | 35.2 (19.7–53.1) | Wesson et al. 2023[7] |
| | Pneumonia | 0–17 years | 11.5 (4.6–22.3) | Prasad et al. 2023[8] |
| | | 18–64 years | 10.7 (9.9–11.5) | Isturiz et al. 2019[6] |
| | | ≥65 years | 9.2 (8.5–9.9) | Isturiz et al. 2019[6] |

Abbreviations: CI – confidence interval; AOM – acute otitis media.

[a] Attributable proportion estimates presented obtained by fitting beta distributions to reported estimates (point estimate and uncertainty intervals) from published studies and estimating the median and 95% confidence limits from five million draws from the fitted distribution.

[b] No published estimate for outpatient adult pneumonia. Inpatient adult pneumonia pneumococcal-attributable percent from Isturiz et al. 2019[6] used for both inpatient and outpatient pneumonia.

[c] No post-PCV13 published estimates available for adult sinusitis. Pneumococcal-attributable proportion of sinusitis cases in adults extrapolated from pediatric published estimate.

**Table S4. Estimated proportions of IPD cases by syndrome[a] from 2018–2019 ABCs data and Prasad et al. 2023[8]**

| Age group | Total N IPD cases | % of IPD cases[a,b] | | |
|---|---|---|---|---|
| | | *Meningitis* | *Bacteremic pneumonia* | *Other bacteremia* |
| Total | 6492 | 8% | 78% | 15% |
| All children | 472 | 17%[c] | 69%[d] | 13%[d] |
| <1 year | 109 | 16%[e] | 71%[d] | 13%[d] |
| 1 year | 81 | 16%[e] | 71%[d] | 13%[d] |
| 2–4 years | 108 | 16%[e] | 71%[d] | 13%[d] |
| 5–17 years | 174 | 20%[e] | 67%[d] | 13%[d] |
| All adults | 6020 | 7%[f] | 78%[g] | 15%[g] |

Abbreviations: IPD – invasive pneumococcal disease; ABCs – Active Bacterial Core surveillance

[a] Categorized into mutually exclusive categories of meningitis, bacteremic pneumonia, and other bacteremia. Any IPD case with meningitis was categorized as meningitis regardless of presence or absence of other syndrome. Bacteremic pneumonia and bacteremia as categorized in ABCs data considered mutually exclusive. We categorized cases without an identified syndrome in ABCs data (~7% of IPD cases) into syndromes based on the distribution of categorized syndromes in ABCs data.

[b] Percents may not sum to 100% due to rounding.

[c] Weighted average of % of IPD cases that are meningitis from Prasad et al. 2023;[8] weighted by age-group specific estimates.

[d] Calculated as the percent of non-meningitis IPD cases (100% – % meningitis) from Prasad et al. 2023[8] multiplied by the proportion of non-meningitis IPD cases belonging to each syndrome based on ABCs data (not stratified by age). Percent of non-meningitis IPD that is bacteremic pneumonia is 84%. Percent of non-meningitis IPD that is other bacteremia is 16%.

[e] From Prasad et al. 2023.[8]

[f] Calculated as (N meningitis cases – N meningitis cases in children) / N meningitis cases. N meningitis cases calculated as Total IPD cases multiplied by the percent of IPD cases that are meningitis (8% x 6492).

[d] Calculated as the percent of non-meningitis IPD cases (100% – % meningitis) as detailed in [f] multiplied by the proportion of non-meningitis IPD cases belonging to each syndrome based on ABCs data.

**Table S5. Health economic burden model inputs**

| Syndrome | Measure[a] | Value by age group | | | | | |
|---|---|---|---|---|---|---|---|
| | | *<2 years* | *2–4 years* | *5–17 years* | *18–49 years* | *50–64 years* | *≥65 years* |
| AOM | % office visits[b] | 87 | 87 | 91 | - | - | - |
| | % ED visits[b] | 13 | 13 | 9 | - | - | - |
| | Median office visit cost, 2022 USD (95% CI)[c] | 49 (14, 120) | 49 (14, 120) | 49 (14, 120) | - | - | - |
| | Median antibiotic prescription cost, 2022 USD (95% CI)[d] | 51 (14, 126) | 56 (16, 137) | 68 (19, 166) | - | - | - |
| | % requiring tympanostomy tube insertion (95% CI)[e] | 12 (8, 17) | 8 (3, 14) | - | - | - | - |
| | Median tympanostomy tube insertion cost, 2022 USD (95% CI)[f] | 3,692 (2,999, 4,483) | 3,692 (2,999, 4,483) | - | - | - | - |
| | QALY decrement per episode[g] | 0.0016 | 0.0016 | 0.0016 | - | - | - |
| | Days of missed work[h] | 0.74 | 0.74 | 0.74 | - | - | - |
| Sinusitis | % office visits[b] | - | - | 97 | 96 | 97 | 98 |
| | % ED visits[b] | - | - | 3 | 4 | 3 | 2 |
| | Median office visit cost, 2022 USD (95% CI)[c] | - | - | 49 (14, 120) | 49 (14, 120) | 49 (14, 120) | 49 (14, 120) |
| | Median antibiotic prescription cost, 2022 USD (95% CI)[i] | - | - | 72 (20, 177) | 104 (29, 255) | 105 (29, 258) | 115 (32, 283) |
| | % requiring surgery[j] | - | - | 2 | 2 | 2 | 2 |
| | Median cost of sinus surgery, 2022 USD (95% CI)[k] | - | - | 1,802 (505, 4,430) | 1,802 (505, 4,430) | 1,802 (505, 4,430) | 1,802 (505, 4,430) |
| | QALY decrement per episode[l] | - | - | 0.0004 | 0.0004 | 0.0004 | 0.0004 |
| | Days of missed work[h] | - | - | 0.74 | 0.74 | 0.74 | 0.74 |
| Outpatient pneumonia | % office visits[b] | 86 | 86 | 87 | 74 | 79 | 83 |
| | % ED visits[b] | 14 | 14 | 13 | 26 | 21 | 17 |
| | Office visit cost, 2022 USD (95% CI)[c] | 49 (14, 120) | 49 (14, 120) | 49 (14, 120) | 78 (22, 192) | 78 (22, 192) | 78 (22, 192) |
| | Median antibiotic prescription cost, 2022 USD (95% CI)[m] | 71 (20, 175) | 88 (25, 217) | 70 (20, 172) | 150 (42, 369) | 176 (49, 432) | 149 (42, 366) |

| | | Value by age group | | | | | |
|---|---|---|---|---|---|---|---|
| Syndrome | Measure[a] | *<2 years* | *2–4 years* | *5–17 years* | *18–49 years* | *50–64 years* | *≥65 years* |
| | QALY decrement per episode[g] | 0.0165 | 0.0165 | 0.0165 | 0.0071 | 0.0071 | 0.0333 |
| | Days of missed work[n] | 7 | 7 | 7 | 7 | 7 | 7 |
| Inpatient pneumonia | Median hospitalization cost, 2022 USD (95% CI)[o] | 11,214 (9,109, 13,622) | 11,214 (9,109, 13,622) | 11,214 (9,109, 13,622) | 9,910 (2,776, 24,373) | 11,376 (3,188, 27,971) | 11,064 (3,100, 27,207) |
| | Hospitalization length of stay, days[p] | 2.0 | 2.0 | 2.0 | 2.9 | 3.4 | 4.1 |
| | % resulting in death[p] | 0.2 | 0.1 | 0.3 | 1.6 | 3.2 | 6.7 |
| | % requiring post-acute care[p] | 0 | 0 | 0 | 0 | 4 | 8 |
| | QALY decrement per episode[g] | 0.0016 | 0.0016 | 0.0016 | 0.0233 | 0.0233 | 0.0493 |
| Mastoiditis | Median hospitalization cost, 2022 USD (95% CI)[q] | 12,589 (3,527, 30,961) | 12,589 (3,527, 30,961) | 12,589 (3,527, 30,961) | - | - | - |
| | Hospitalization length of stay, days[q] | 4.3 | 4.3 | 4.3 | - | - | - |
| | QALY decrement per episode[r] | 0.0016 | 0.0016 | 0.0016 | - | - | - |
| Meningitis | % of IPD that is meningitis[s] | 16.0 | 16.0 | 20.0 | 6.9 | 6.9 | 6.9 |
| | % hospitalized[t] | 100.0 | 100.0 | 100.0 | 100.0 | 100.0 | 100.0 |
| | Median hospitalization cost, 2022 USD (95% CI)[u,v] | 26,275 (21,343, 31,915) | 26,275 (21,343, 31,915) | 26,275 (21,343, 31,915) | 19,601 (5,491, 48,204) | 24,810 (6,952, 61,011) | 19,293 (5,405, 47,449) |
| | Hospitalization length of stay, days[w] | 8.1 | 4.0 | 4.0 | 5.0 | 6.7 | 6.3 |
| | QALY decrement per episode[x] | 0.003 | 0.003 | 0.003 | 0.0431 | 0.0431 | 0.0431 |
| | % resulting in death[y] | 2.0 | 4.5 | 3.9 | 7.5 | 9.7 | 17.4 |
| | % resulting in disability[y] | 7.0 | 7.0 | 11.0 | 22.0 | 22.0 | 22.0 |
| | QALY decrement from disability[z] | 0.2100 | 0.2100 | 0.2100 | 0.5865 | 0.5865 | 0.5865 |
| | % resulting in deafness[y] | 9.0 | 9.0 | 9.0 | 10.0 | 10.0 | 10.0 |

| Syndrome | Measure[a] | Value by age group *<2 years* | *2–4 years* | *5–17 years* | *18–49 years* | *50–64 years* | *≥65 years* |
|---|---|---|---|---|---|---|---|
| | QALY decrement from deafness[z] | 0.1500 | 0.1500 | 0.1500 | 0.3650 | 0.3650 | 0.3650 |
| | % resulting in mild/moderate hearing loss (95% CI)[aa] | 7.8 (6.1, 12.1) | 7.8 (6.1, 12.1) | 7.8 (6.1, 12.1) | 7.8 (6.1, 12.1) | 7.8 (6.1, 12.1) | 7.8 (6.1, 12.1) |
| | QALY decrement from mild/moderate hearing loss[bb] | 0.027 | 0.027 | 0.027 | 0.027 | 0.027 | 0.027 |
| | % requiring post-acute care[cc] | 7.6 | 7.6 | 7.6 | 7.6 | 7.6 | 7.6 |
| Non-meningitis IPD | % of IPD that is bacteremic pneumonia[s] | 70.7 | 70.7 | 67.3 | 78.3 | 78.3 | 78.3 |
| | % of IPD that is other bacteremia[s] | 13.3 | 13.3 | 12.7 | 14.8 | 14.8 | 14.8 |
| | % hospitalized[t] | 100.0 | 100.0 | 100.0 | 100.0 | 100.0 | 100.0 |
| | Hospitalization cost, 2022 USD (95% CI)[dd,v] | 19,630 (15,941, 23,849) | 19,630 (15,941, 23,849) | 19,630 (15,941, 23,849) | 19,601 (5,491, 48,204) | 24,810 (6,952, 61,011) | 19,293 (5,405, 47,449) |
| | Hospitalization length of stay, days[w] | 5.7 | 5.7 | 5.7 | 7.4 | 7.4 | 7.4 |
| | QALY decrement per episode, bacteremic pneumonia[ee] | 0.0041 | 0.0041 | 0.0041 | 0.0233 | 0.0233 | 0.0493 |
| | QALY decrement per episode, other bacteremia[ff] | 0.0041 | 0.0041 | 0.0041 | 0.0373 | 0.0373 | 0.0373 |
| | % resulting in death[t] | 5.0 | 5.0 | 5.0 | 20.0 | 20.0 | 20.0 |
| | % bacteremic pneumonia requiring post-acute care[gg] | 0 | 0 | 0 | 0 | 4 | 8 |
| | % other bacteremia requiring post-acute care[gg] | 0 | 0 | 0 | 0 | 0 | 0 |
| All | Median emergency department cost, 2022 USD (95% CI)[hh] | 222 (62, 547) | 222 (62, 547) | 222 (62, 547) | 222 (62, 547) | 222 (62, 547) | 222 (62, 547) |
| | Value of day of missed work, 2022 USD[ii] | 161 | 161 | 161 | 161 | 152 | 35 |

| Syndrome | Measure[a] | Value by age group <2 years | 2–4 years | 5–17 years | 18–49 years | 50–64 years | ≥65 years |
|---|---|---|---|---|---|---|---|
| | Median out of pocket costs, outpatient syndromes, 2022 USD (95% CI)[jj] | 21 (6, 52) | 21 (6, 52) | 21 (6, 52) | 21 (6, 52) | 21 (6, 52) | 21 (6, 52) |
| | Median post-acute care cost, 2022 USD (95% CI)[kk] | 3,932 (1,102, 9,667) | 3,932 (1,102, 9,667) | 3,932 (1,102, 9,667) | 3,932 (1,102, 9,667) | 3,932 (1,102, 9,667) | 3,932 (1,102, 9,667) |
| | US GDP per capita, 2022 USD[ll] | 76,399 | 76,399 | 76,399 | 76,399 | 76,399 | 76,399 |

Abbreviations: AOM – acute otitis media; ED – emergency department; USD – United States dollars; CI – confidence interval; IPD – invasive pneumococcal disease; QALY – quality adjusted life-year; US – United States; GDP – gross domestic product

[a] Uncertainty (95% CI) estimated for cost parameters. For all other parameters, uncertainty only included if reported in original data source.

[b] Percent of visits in emergency department versus office visit settings calculated from Table 2 in Huang et al. 2011.[9]

[c] Obtained by fitting a gamma distribution to estimates from Huang et al. 2011[9] of $42 (AOM, sinusitis, pediatric outpatient pneumonia) and $67 (adult outpatient pneumonia) in 2007 USD and assuming standard deviations of 50% of the mean (variance not reported in original source), scaled to 2022 USD.

[d] Obtained by fitting a gamma distribution to estimates from Huang et al. 2011[9] of $44 (<2 years), $48 (2–4 years), and $58 (5–17 years) in 2007 USD and assuming standard deviations of 50% of the mean (variance not reported), scaled to 2022 USD.

[e] Percent requiring tympanostomy tube insertion obtained by fitting reported values from Prasad et al. 2023[8] of 12% (95% CI 6%, 15%; <2 years) and 8% (95% CI 4%,15%; 2–4 years) to a beta distribution.

[f] Obtained by fitting a gamma distribution to the reported value of $3,449 (95% CI $2,759, $4,138) in 2021 USD from Prasad et al. 2023,[8] scaled to 2022 USD.

[g] QALY decrements from Tang et al. 2022.[10]

[h] Parental missed work due to AOM. Calculated as 5.9 hours of work missed per episode (from Capra et al. 2000[11]) divided by 8 hours of work per day, based assumption of 40-hour work week. Work missed for sinusitis assumed to be similar to AOM for adults and children.

[i] Obtained by fitting a gamma distribution to estimates from Huang et al. 2011[9] of $62 (5–17 years), $89 (18–49 years), $90 (50–64 years), and $99 (≥65 years) in 2007 USD and assuming standard deviations of 50% of the mean (variance not reported), scaled to 2022 USD.

[j] Percent of sinusitis cases requiring surgery from Huang et al. 2011.[9]

[k] Obtained by fitting a gamma distribution to estimate of $1,548 in 2007 USD from Huang et al. 2011[9] and assuming a standard deviation of 50% of the mean (variance not reported), scaled to 2022 USD.

[l] Estimated as the product of sinusitis duration (7 days[12])/365 days and sinusitis disutility of 0.02 from Melse et al. 2000.[13]

[m] Obtained by fitting a gamma distribution to estimates from Huang et al. 2011[9] of $61 (<2 years), $76 (2–4 years), $60 (5–17 years), $129 (18–49 years), $151 (50–64 years), and $128 (≥65 years) in 2007 USD and assuming standard deviations of 50% of the mean (variance not reported), scaled to 2022 USD.
[n] Based on median return to work time for outpatient pneumonia cases from Fine et al. 1999.[14]
[o] Pneumonia hospitalization cost for children 0–17 years obtained by fitting a gamma distribution to estimate of $10,475 (95% CI $8,380–$12,570) in 2021 USD from Prasad et al. 2023[8] scaled to 2022 USD. Estimates for adult hospitalization costs obtained by fitting gamma distributions to estimates from Rubin et al. 2010[15] in 2008 USD: $8,843 (18–49 years; population weighted average of 18–34 and 35–49 year estimates), $10,148 (50–64 years), and $9,872 (≥65 years) assuming standard deviations of 50% of the mean (variance not reported), scaled to 2022 USD.
[p] Inpatient pneumonia length of stay, percent resulting in death, and percent requiring post-acute cure from Huang et al. 2011.[9]
[q] Mastoiditis median hospitalization cost and length of stay from Acevedo et al. 2009.[16] Hospitalization costs obtained by fitting a gamma distribution to the reported median hospitalization cost of $9,600 in 2003 USD, assuming a standard deviation of 50% of the mean (variance not reported), scaled to 2022 USD. Length of stay value calculated as the population-weighted average of all length of stay values from Table 2 in Acevedo et al. 2009.
[r] Mastoiditis QALY decrement values not available, conservatively assumed to be equivalent to AOM, consistent with previous studies.[17] QALY decrement value from Tang et al. 2022.[10]
[s] Proportion of IPD cases that are meningitis and non-meningitis (bacteremia and bacteremic pneumonia) in children 0–17 years from Prasad et al. 2023.[88] Proportion of IPD cases that are meningitis and non-meningitis in adults ≥18 years estimated based on 2018–2019 CDC Active Bacterial Core Surveillance data and Prasad et al. 2023.[8] Calculations detailed in **Table S4**.
[t] Proportion of IPD cases hospitalized and resulting in death from Huang et al. 2011.[9]
[u] Meningitis hospitalization cost for <2 years, 2–4 years, and 5–17 years obtained by fitting a gamma distribution to $24,544 (95% CI 19,635–29,453) in 2021 USD from Prasad et al. 2023,[8] scaled to 2022 USD.
[v] All IPD hospitalization costs for adults 18–49 years obtained by fitting a gamma distribution to estimate of $17,490.73 (population-weighted average of 18–34 and 35–49 year estimates) in 2008 USD from Rubin et al. 2010,[15] assuming a standard deviation of 50% of the mean (variance not reported), scaled to 2022 USD. IPD hospitalization costs for 50–64 years and ≥65 years obtained by fitting gamma distribution to reported means of USD 2008 $22,135 and $17,216, respectively, from Rubin et al. 2010 assuming standard deviations of 50% of the reported means (variance not reported), scaled to 2022 USD.
[w] Hospitalization length of stay from Huang et al. 2011;[9] typical resolution value used. For non-meningitis IPD, bacteremia length of stay values used for other bacteremia and bacteremic pneumonia.
[x] QALY decrements from Tang et al. 2022. Values for meningitis without sequalae used for most conservative estimate as QALY decrements from sequelae considered separately and for consistency between adults and children.
[y] Percent of meningitis cases resulting in disability and deafness among children 0–17 years from Prasad et al. 2023.[8] Percent of meningitis cases resulting in disability and deafness among adults ≥18 years from Huang et al. 2011.[9] Point estimates only used in analyses; uncertainty not reported in all studies.
[z] QALY decrements for disability and deafness from Tang et al. 2022,[10] median values. Decrements due to disability and deafness not provided for ≥65 years; we assumed constant rates for all adults and used estimated value from adults 19–64 years.

[aa] Percent of meningitis cases resulting in mild to moderate hearing loss from Edmond et al. 2010.[18]
[bb] QALY decrements for hearing loss from Tordrup et al. 2022,[19] moderate hearing loss decrement used.
[cc] Percent of meningitis cases requiring post-acute care from Ellis et al. 2020.[20]
[dd] Hospitalization cost for <2 years, 2–4 years, and 5–17 years obtained by fitting a gamma distribution to estimate of $18,339 (95% CI $14,671, $22,007) in 2021 USD from Prasad et al. 2023,[8] scaled to 2022 USD.
[ee] From Tang et al. 2022.[10] Where both inpatient pneumonia and bacteremia QALY values provided, we used only the value with the greatest decrement to capture the driver of QALY losses without double counting QALY losses due to a single condition.
[ff] From Tang et al. 2022.[10] We assumed QALY decrements for bacteremia were consistent across all adults and used the value for adults ≥65 years for all adults as no value provided for adults <65 years.
[gg] Percent of non-meningitis IPD cases requiring post-acute care from Huang et al. 2011.[9]
[hh] Obtained by fitting a gamma distribution to estimate from Huang et al. 2011[9] of $191 in 2007 USD assuming a standard deviation of 50% of the mean (variance not reported), scaled to 2022 USD.
[ii] Scaled from original 2007 USD values of $114 in 0–49 years, $108 in 50–64 years, and $25 in ≥65 years from Huang et al. 2011.[9]
[jj] Obtained by fitting a gamma distribution to estimate of $18 in 2007 USD from Huang et al. 2011[9] and assuming standard deviations of 50% of the mean (variance not reported), scaled to 2022 USD.
[kk] Obtained by fitting a gamma distribution to estimate of $3,378 in 2007 USD from Huang et al. 2011[9] and assuming standard deviations of 50% of the mean (variance not reported), scaled to 2022 USD.
[ll] US GDP per capita in 2022 from World Bank data.[21]

**Table S6. Sensitivity analyses**

| Scenario compared with primary analysis | Sensitivity analysis value(s)[a] | Sensitivity analysis values source | Change in cost estimate compared to primary analysis, in millions, 2022 USD (95% CI) | |
|---|---|---|---|---|
| | | | *PCV24-additional serotypes* | *PCV31-additional serotypes* |
| Lower probability of hospitalization for IPD[b] | Meningitis <5 years: 1.0; meningitis ≥5 years: 0.97; non-meningitis IPD <5 years: 0.82; non-meningitis IPD ≥5 years: 0.83 | Prasad et al. 2023[8] | -7.4 (-13.3, -3.8) | -34.6 (-59.7, -18.4) |
| Greater probability of disability from meningitis in children | <2 years: 0.24; 2–17 years: 0.37 | Huang et al. 2011[9] | 1.8 (1.3, 2.8) | 11.7 (9.9, 13.9) |
| Varying probability of ED visits for outpatient conditions | 0.10 | Andrejko et al. 2022[22] | 2.8 (0.0, 9.3) | 34.5 (-0.3, 111.8) |
| Lower pneumococcal-attributable percent of sinusitis cases | 11.8% (1.0%, 22.7%) | King et al. 2024[c1] | -77.9 (-138.5, -31.1) | -909.8 (-1,586.0, -366.2) |
| Lower pneumococcal-attributable percent of outpatient pediatric pneumonia | 11.8% (1.0%, 22.7%) | King et al. 2024[c1] | -48.7 (-67.9, -34.4) | -321.0 (-498.4, -183.6) |
| Lower cost of non-bacteremic pneumonia hospitalization | 0–17 years: $2,581 ($723, $6,348); 18–49 years: $4,050 ($1,135, $9,959); 50–64 years: $4,736 ($1,327, $11,648); ≥65 years: $4,989 ($1,398, $12,269)[d] | Huang et al. 2011[9] | -72.8 (-185.5, 3.1) | -389.1 (-884.8, -38.0) |

| Scenario compared with primary analysis | Sensitivity analysis value(s)[a] | Sensitivity analysis values source | Change in cost estimate compared to primary analysis, in millions, 2022 USD (95% CI) | |
|---|---|---|---|---|
| | | | *PCV24-additional serotypes* | *PCV31-additional serotypes* |
| Higher cost of non-bacteremic pneumonia hospitalization | 0–17 years: $38032 ($10,655, $93,530); 18–49 years: $22,993 ($6,447, $56,526); 50–64 years: $24,749 ($6,933, $60,870); ≥65 years: $16,387 ($4,592, $40,297)[e] | Huang et al. 2023[23] | 116.4 (-29.4, 298.2) | 716.4 (-28.3, 1,688.6) |
| Lower cost of meningitis hospitalization | <2 years: $17,690 ($4,955, $43,508); 2–17 years: $8,768 ($2,457, $21,563); 18–49 years: $10,945 ($3,066, $26,919); 50–64 years: $14,646 ($4,103, $36,020); ≥65 years: $11,593 ($3,246, $28,519)[f] | Huang et al. 2011[9] | -1.5 (-4.0, 0.5) | -7.1 (-18.5, 2.2) |
| Higher cost of meningitis hospitalization | 0–17 years: $58,261 ($16,328, $143,259); 18–49 years: $51,347 ($14,383, $126,285); 50–64 years: $52,449 ($14,699, $128,966); ≥65 years: $25,129 ($7,040, $61,797)[g] | Huang et al. 2023[23] | 3.3 (-0.4, 8.1) | 16.2 (-1.1, 38.2) |
| Lower cost of non-meningitis hospitalization | 0–17 years: $10,992 ($3,080, $27,029); 18–64 years: $12,866 ($3,604, $31,642); ≥65 years: $10,873 ($3,046, $26,740)[h] | Huang et al. 2011[9] | -18.7 (-51.0, 7.6) | -87.3 (-232.7, 33.3) |

| Scenario compared with primary analysis | Sensitivity analysis value(s)[a] | Sensitivity analysis values source | Change in cost estimate compared to primary analysis, in millions, 2022 USD (95% CI) | |
|---|---|---|---|---|
| | | | *PCV24-additional serotypes* | *PCV31-additional serotypes* |
| Higher cost of non-meningitis hospitalization | 0–17 years, bacteremic pneumonia: \$52,333 (\$14,662, \$143,128,698); 0–17 years, other bacteremia: \$41,769 (\$11,699, \$102,732); 18–49 years: \$51,347 (\$14,383, \$126,285); 50–64 years: \$52,449 (\$14,699, \$128,966); ≥65 years: \$25,129 (\$7,040, \$61,797)[i] | Huang et al. 2023[23] | 37.9 (-9.2, 101.1) | 108.1 (-37.1, 459.3) |

[a] Stratified by age group, where applicable. Costs are in 2022 USD.
[b] Prasad et al. estimated probability of hospitalization for meningitis and non-meningitis IPD cases for children 5–20 years. We extrapolated that estimate to all individuals >5 years of age.
[c] Primary analysis used attributable proportion from King et al. 2024[1] vaccine probe approach (primary analysis) while sensitivity analyses used attributable proportion from King et al. 2024[1] differential carriage approach.
[d] Obtained by fitting gamma distributions to estimates from Huang et al. 2011[9] of \$2,218 (0–17 years), \$3,480 (18–49 years), \$4,070 (50–64 years), and \$4,287 (≥65 years) in 2007 USD and assuming standard deviations of 50% of the mean (variance not reported), scaled to 2022 USD.
[e] Obtained by fitting gamma distributions to estimates from Huang et al. 2023[23] of \$42,708 (0–17 years), \$25,814 (18–49 years), \$27,797 (50–64 years), and \$18,400 (≥65 years) in 2021 USD and assuming standard deviations of 50% of the mean (variance not reported), scaled to 2022 USD.
[f] Obtained by fitting gamma distributions to estimates from Huang et al. 2011[9] of \$15,202 (<2 years), \$7,536 (2–17 years), \$9,406 (18–49 years), \$12,585 (50–64 years), and \$9,965 (≥65 years) in 2007 USD and assuming standard deviations of 50% of the mean (variance not reported), scaled to 2022 USD.
[g] Obtained by fitting gamma distributions to estimates from Huang et al. 2023[23] of \$65,419 (bacteremia 0–17 years), \$57,657 (18–49 years), \$58,890 (50–64 years), and \$28,217 (≥65 years) in 2021 USD and assuming standard deviations of 50% of the mean (variance not reported), scaled to 2022 USD.

[h] Obtained by fitting gamma distributions to estimates from Huang et al. 2011[9] of $9,444 (0–17 years), $11,055 (18–64 years), and $9,344 (≥65 years) in 2007 USD and assuming standard deviations of 50% of the mean (variance not reported), scaled to 2022 USD. Bacteremia values used for all non-meningitis IPD cases.
[i] Obtained by fitting gamma distributions to estimates from Huang et al. 2023[23] of $58,774 (bacteremic pneumonia, 0–17 years), $46,909 (other bacteremia, 0–17 years), $57,657 (all IPD, 18–49 years), $58,890 (all IPD, 50–64 years), and $28,217 (all IPD, ≥65 years) in 2021 USD and assuming standard deviations of 50% of the mean (variance not reported), scaled to 2022 USD.

**Table S7. Estimated incidence[a] and national annual burdens[b] attributable to non-invasive pneumococcal infections (all serotypes)**

| Condition | Age group | Outpatient visits | | Antibiotic prescriptions | | Hospitalizations | |
|---|---|---|---|---|---|---|---|
| | | *Incidence/ 1000 person-years (95% CI)* | *No. in thousands (95% CI)* | *Incidence/ 1000 person-years (95% CI)* | *No. in thousands (95% CI)* | *Incidence/ 100,000 person-years (95% CI)* | *No. in thousands (95% CI)* |
| AOM | <2 years | 116.2 (79.4, 164.5) | 883.9 (603.7, 1,251.3) | 83.1 (54.8, 121.1) | 632.1 (416.4, 920.9) | 1.2 (0.7, 1.9) | 0.0 (0.0, 0.1) |
| | 2–4 years | 68.3 (47.3, 95.7) | 817.5 (566.0, 1,144.8) | 62.2 (44.3, 85.3) | 744.7 (529.5, 1,020.7) | 0.6 (0.3, 0.9) | 0.0 (0.0, 0.1) |
| | 5-17 years | 12.0 (7.7, 17.9) | 643.8 (413.9, 955.4) | 12.3 (9.4, 15.7) | 656.0 (505.0, 840.8) | 0.6 (0.3, 0.9) | 0.3 (0.2, 0.5) |
| | Total[c] | 32.3 (25.2, 41.0) | 2,362.5 (1,839.7, 2,996.9) | 28.0 (22.3, 34.8) | 2,043.6 (1,630.9, 2,540.3) | 0.6 (0.3, 1.0) | 0.5 (0.3, 0.7) |
| Sinusitis | 5–17 years | 22.7 (12.7, 37.0) | 1,217.3 (678.5 1,978.3) | 20.9 (11.6, 33.8) | 1,115.9 (622.7, 1,811.1) | - | - |
| | 18–49 years | 37.2 (23.0, 52.6) | 5,147.8 (3,180.4, 7,271.4) | 25.9 (16.0, 36.8) | 3,585.6 (2,208.2, 5,093.2) | - | - |
| | 50–64 years | 41.4 (25.6, 58.6) | 2,605.0 (1,607.8, 3,684.3) | 27.5 (16.9, 39.2) | 1,732.3 (1,065.6, 2,465.9) | - | - |
| | ≥65 years | 19.7 (11.7, 29.6) | 1,065.6 (633.9, 1,601.5) | 11.2 (6.4, 17.7) | 607.7 (348.4, 957.5) | - | - |
| | Total[c] | 32.6 (20.2, 45.9) | 10,074.6 (6,231.7, 14,181.7) | 22.9 (14.1, 32.4) | 7,076.4 (4,368.7, 10,007.6) | - | - |
| Pneumonia | <2 years | 12.2 (6.2, 22.1) | 92.7 (46.9, 167.9) | 9.2 (5.3, 14.9) | 69.7 (39.9, 113.6) | 118.9 (76.1, 169.6) | 9.0 (5.8, 12.9) |
| | 2–4 years | 7.7 (4.0, 13.9) | 92.4 (47.3 166.1) | 7.5 (4.5, 11.7) | 89.8 (53.6, 140.4) | 85.1 (54.4, 121.5) | 10.2 (6.5, 14.5) |

| Condition | Age group | Outpatient visits | | Antibiotic prescriptions | | Hospitalizations | |
|---|---|---|---|---|---|---|---|
| | | *Incidence/ 1000 person-years (95% CI)* | *No. in thousands (95% CI)* | *Incidence/ 1000 person-years (95% CI)* | *No. in thousands (95% CI)* | *Incidence/ 100,000 person-years (95% CI)* | *No. in thousands (95% CI)* |
| | 5–17 years | 2.7 (1.4, 4.9) | 144.9 (73.5, 262.6) | 2.2 (1.3, 3.5) | 117.4 (68.5, 187.9) | 18.6 (11.9, 26.6) | 10.0 (6.4, 14.2) |
| | 18–49 years | 0.6 (0.4, 0.7) | 79.4 (61.7, 100.5) | 0.3 (0.2, 0.4) | 43.3 (30.0, 60.1) | 17.8 (16.3, 19.3) | 24.6 (22.6, 26.7) |
| | 50–64 years | 0.6 (0.4, 0.7) | 36.1 (28.0, 45.7) | 0.3 (0.2, 0.4) | 19.7 (13.7, 27.3) | 49.4 (45.2, 53.9) | 31.1 (28.4, 33.9) |
| | ≥65 years | 1.8 (1.3, 2.4) | 95.8 (69.6, 128.3) | 0.8 (0.5, 1.1) | 41.6 (26.7, 61.3) | 176.0 (162.2, 190.4) | 95.1 (87.7, 102.9) |
| | Total[c] | 1.1 (0.8, 1.5) | 358.6 (274.8, 481.9) | 0.7 (0.5, 0.9) | 223.6 (166.8, 299.0) | 54.9 (50.6, 59.5) | 180.1 (166.1, 195.4) |

[a] Estimated incidence rates for outpatient visits and antibiotic prescriptions reported per 1,000 person-years. Estimated incidence rates for hospitalizations reported per 100,000 person-years.
[b] Estimated using 2019 bridged-race census (Vintage 2020)[24] values for each age group.
[c] Component values may not sum exactly to totals due to rounding and uncertainty propagation methods.

**Table S8. Estimated incidence of invasive pneumococcal disease attributable to PCV24- and PCV31-additional serotypes,[a] 2019**

| | Incidence per 100,000 person-years (95% CI) | | |
|---|---|---|---|
| **Age group[b]** | *All serotypes[c]* | *PCV24-additional serotypes* | *PCV31-additional serotypes* |
| <2 years | 12.0 | 0.3 (0.1, 0.8) | 3.2 (2.4, 4.3) |
| 2–4 years | 4.3 | 0.1 (0.0, 0.3) | 1.1 (0.9, 1.5) |
| 5–17 years | 1.4 | 0.1 (0.1, 0.1) | 0.4 (0.4, 0.5) |
| 18–49 years | 4.4 | 0.3 (0.3, 0.4) | 1.4 (1.3, 1.5) |
| 50–64 years | 15.6 | 1.1 (0.9, 1.3) | 4.9 (4.6, 5.2) |
| ≥65 years | 23.7 | 1.6 (1.4, 2.0) | 7.4 (7.0, 7.9) |
| Total | 9.4[d] | 0.6 (0.5, 0.8) | 2.9 (2.7, 3.1) |

Abbreviations: PCV – pneumococcal conjugate vaccine; IPD – invasive pneumococcal disease; CI – confidence interval.

[a] PCV24-additional serotypes (not included in PCV20): 2, 9N, 17F, 20B. PCV31-additional serotypes: 2, 9N, 17F, 20B, 7C, 15A, 16F, 23A, 23B, 31, 35B.

[b] Where study age groups contained multiple ABCs-defined age groups, study age group incidence was estimated as the population-weighted average of component ABCs-defined age group incidence values.

[c] From CDC Active Bacterial Core Surveillance (ABCs) data, no uncertainty reported.

**[d]** Calculated as estimated national N cases divided by 2019 census multiplied by 100,000; differs from ABCs estimate of 9.2 cases/100,000 persons.[25]

**Table S9. Estimated incidence of healthcare utilization attributable to non-invasive PCV24- and PCV31-additional serotype[a] disease**

| | | Outpatient visits per 1,000 person-years (95% CI) | | Antibiotic prescriptions per 1,000 person-years (95% CI) | | Hospitalizations per 100,000 person-years (95% CI) | |
|---|---|---|---|---|---|---|---|
| **Condition** | **Age group** | *PCV24-additional serotypes[a]* | *PCV31-additional serotypes[a]* | *PCV24-additional serotypes[a]* | *PCV31-additional serotypes[a]* | *PCV24-additional serotypes[a]* | *PCV31-additional serotypes[a]* |
| AOM[b] | <2 years | 3.5 (2.3–5.1) | 42.6 (29.0–60.4) | 2.5 (1.6–3.8) | 30.5 (20.0–44.5) | 0.0 (0.0–0.0) | 0.4 (0.2–0.7) |
| | 2–4 years | 2.0 (1.4–3.0) | 25.0 (17.3–35.1) | 1.9 (1.3–2.7) | 22.8 (16.2–31.3) | 0.0 (0.0–0.0) | 0.2 (0.1–0.3) |
| | 5–17 years | 0.4 (0.2–0.6) | 4.4 (2.8–6.6) | 0.4 (0.3–0.5) | 4.5 (3.5–5.8) | 0.0 (0.0–0.0) | 0.2 (0.1–0.3) |
| | All | 1.0 (0.7–1.3) | 11.8 (9.2–15.1) | 0.8 (0.6–1.1) | 10.2 (8.2–12.8) | 0.0 (0.0–0.0) | 0.2 (0.1–0.3) |
| Sinusitis | 5–17 years | 0.7 (0.4–1.2) | 8.1 (4.5–13.2) | 0.6 (0.3–1.1) | 7.4 (4.1–12.1) | - | - |
| | 18–49 years | 1.1 (0.7–1.7) | 13.3 (8.2–18.8) | 0.8 (0.5–1.2) | 9.2 (5.7–13.2) | - | - |
| | 50–64 years | 1.3 (0.8–1.8) | 14.8 (9.1–20.9) | 0.8 (0.5–1.2) | 9.8 (6.0–14.0) | - | - |
| | ≥65 years | 0.6 (0.4–0.9) | 7.0 (4.2–10.6) | 0.3 (0.2–0.6) | 4.0 (2.3–6.3) | - | - |
| | All | 1.0 (0.6–1.4) | 11.6 (7.2–16.4) | 0.7 (0.4–1.0) | 8.2 (5.0–11.6) | - | - |
| Pneumonia | 0–2 years | 0.4 (0.2–0.7) | 4.3 (2.2–7.9) | 0.3 (0.2–0.5) | 3.3 (1.9–5.3) | 3.6 (2.3–5.3) | 42.4 (27.1–60.6) |
| | 2–4 years | 0.2 (0.1–0.4) | 2.8 (1.4–5.0) | 0.2 (0.1–0.4) | 2.7 (1.6–4.2) | 2.6 (1.6–3.8) | 30.3 (19.4–43.4) |
| | 5–17 years | 0.1 (0.0–0.2) | 1.0 (0.5–1.8) | 0.1 (0.0–0.1) | 0.8 (0.5–1.3) | 0.6 (0.4–0.8) | 6.6 (4.2–9.5) |
| | 18–49 years | 0.0 (0.0–0.1) | 0.2 (0.1–0.2) | 0.0 (0.0–0.0) | 0.1 (0.1–0.1) | 1.2 (1.0–1.5) | 5.6 (5.0–6.2) |
| | 50–64 years | 0.0 (0.0–0.1) | 0.2 (0.1–0.2) | 0.0 (0.0–0.0) | 0.1 (0.1–0.1) | 3.4 (2.8–4.3) | 15.5 (13.9–17.3) |
| | ≥65 years | 0.1 (0.1–0.2) | 0.6 (0.4–0.8) | 0.1 (0.0–0.1) | 0.2 (0.2–0.4) | 12.0 (10.0–15.3) | 55.2 (49.9–61.1) |
| | All | 0.1 (0.1–0.1) | 0.6 (0.4–0.8) | 0.0 (0.0–0.1) | 0.4 (0.3–0.6) | 3.4 (2.9–4.3) | 17.6 (15.9–19.5) |

| | | Outpatient visits per 1,000 person-years (95% CI) | | Antibiotic prescriptions per 1,000 person-years (95% CI) | | Hospitalizations per 100,000 person-years (95% CI) | |
|---|---|---|---|---|---|---|---|
| **Condition** | **Age group** | *PCV24-additional serotypes*[a] | *PCV31-additional serotypes*[a] | *PCV24-additional serotypes*[a] | *PCV31-additional serotypes*[a] | *PCV24-additional serotypes*[a] | *PCV31-additional serotypes*[a] |
| All | 0–2 years | 3.9 (2.6–5.6) | 47.1 (33.3–65.2) | 2.8 (1.8–4.1) | 33.8 (23.2–47.9) | 3.6 (2.3–5.3) | 42.8 (27.5–61.0) |
| | 2–4 years | 2.3 (1.6–3.3) | 27.9 (20.0–38.2) | 2.1 (1.5–2.9) | 25.5 (18.8–34.2) | 2.6 (1.6–3.8) | 30.5 (19.6–43.6) |
| | 5–17 years | 1.1 (0.8–1.7) | 13.6 (9.5–19.1) | 1.1 (0.8–1.5) | 12.8 (9.2–17.6) | 0.6 (0.4–0.8) | 6.8 (4.4–9.7) |
| | 18–49 years | 0.9 (0.6 –1.3) | 10.6 (6.6–15.0) | 0.6 (0.4–0.9) | 7.4 (4.6–10.4) | 1.2 (1.0–1.5) | 5.6 (5.0–6.2) |
| | 50–64 years | 1.3 (0.8–1.9) | 14.9 (9.3–21.1) | 0.9 (0.5 –1.3) | 9.9 (6.1–14.1) | 3.4 (2.8–4.3) | 15.5 (13.9–17.3) |
| | ≥65 years | 0.7 (0.5–1.1) | 7.6 (4.7–11.2) | 0.4 (0.2–0.6) | 4.3 (2.5–6.6) | 12.0 (10.0–15.3) | 55.2 (49.9–61.1) |
| | All | 1.2 (0.8–1.7) | 14.2 (9.9–18.7) | 0.9 (0.6–1.2) | 10.4 (7.4–13.6) | 3.4 (2.9–4.3) | 17.6 (15.9–19.5) |

Abbreviations: PCV – pneumococcal conjugate vaccine; CI – confidence interval; AOM – acute otitis media.
[a] PCV24-additional serotypes (not included in PCV20): 2, 9N, 17F, 20B. PCV31-additional serotypes: 2, 9N, 17F, 20B, 7C, 15A, 16F, 23A, 23B, 31, 35B.
[b] AOM-related hospitalizations are for mastoiditis.

**Table S10. Annual inpatient health economic burden attributable to non-invasive PCV24-additional serotype[a] pneumococcal disease**

| | | Costs in millions, 2022 USD (95% CI) | | | | | | | |
|---|---|---|---|---|---|---|---|---|---|
| | | **Direct medical costs** | | | **Non-medical costs** | | | | ***Total*** |
| **Condition** | **Age group** | *Hospitalization* | *Post-acute care* | *All medical[b]* | *Missed work* | *QALYs lost[c]* | *YLL due to death* | *All non-medical* | |
| Mastoiditis | <2 years | 0.0 (0.0, 0.0) | - | 0.0 (0.0, 0.0) | 0.0 (0.0, 0.0) | 0.0 (0.0, 0.0) | - | 0.0 (0.0, 0.0) | 0.0 (0.0, 0.0) |
| | 2–4 years | 0.0 (0.0, 0.0) | - | 0.0 (0.0, 0.0) | 0.0 (0.0, 0.0) | 0.0 (0.0, 0.0) | - | 0.0 (0.0, 0.0) | 0.0 (0.0, 0.0) |
| | 5–17 years | 0.0 (0.0, 0.1) | - | 0.0 (0.0, 0.1) | 0.0 (0.0, 0.0) | 0.0 (0.0, 0.0) | - | 0.0 (0.0, 0.0) | 0.0 (0.0, 0.1) |
| | Total[b] | 0.0 (0.0, 0.1) | - | 0.0 (0.0, 0.1) | 0.0 (0.0, 0.0) | 0.0 (0.0, 0.0) | - | 0.0 (0.0, 0.0) | 0.0 (0.0, 0.1) |
| Pneumonia | <2 years | 3.1 (1.9, 4.8) | - | 3.1 (1.9, 4.8) | 0.1 (0.1, 0.1) | 0.0 (0.0, 0.0) | 1.3 (0.8, 1.9) | 1.4 (0.9, 2.1) | 4.5 (2.8, 6.8) |
| | 2–4 years | 3.5 (2.1, 5.4) | - | 3.5 (2.1, 5.4) | 0.1 (0.1, 0.1) | 0.0 (0.0, 0.1) | 0.7 (0.4, 1.1) | 0.9 (0.5, 1.3) | 4.3 (2.6, 6.6) |
| | 5–17 years | 3.4 (2.0, 5.3) | - | 3.4 (2.0, 5.3) | 0.1 (0.1, 0.1) | 0.0 (0.0, 0.1) | 2.0 (1.3, 3.0) | 2.2 (1.4, 3.2) | 5.6 (3.4, 8.4) |
| | 18–49 years | 16.6 (4.6, 42.1) | - | 16.6 (4.6, 42.1) | 0.8 (0.7, 1.0) | 2.9 (2.5, 3.7) | 51.5 (43.2, 66.0) | 55.2 (46.4, 70.8) | 72.5 (55.4, 103.9) |
| | 50–64 years | 24.2 (6.7, 61.1) | 0.3 (0.1, 0.8) | 24.5 (7.0, 61.5) | 1.1 (0.9, 1.4) | 3.6 (3.0, 4.7) | 94.4 (79.0, 121.1) | 99.1 (83.0, 127.2) | 124.7 (97.3, 173.8) |
| | ≥65 years | 71.9 (19.9, 181.7) | 1.9 (0.5, 4.8) | 74.0 (21.9, 184.0) | 0.9 (0.8, 1.2) | 22.7 (19.1, 29.1) | 374.4 (314.6, 479.5) | 398.0 (334.5, 509.8) | 475.8 (381.0, 641.3) |
| | Total[b] | 127.7 (64.4, 245.1) | 2.2 (0.6, 5.6) | 130.2 (66.4, 247.8) | 3.1 (2.6, 3.9) | 29.4 (24.9, 37.5) | 524.3 (444.4, 668.9) | 556.8 (471.9, 710.4) | 690.5 (566.5, 902.4) |
| All[b] | <2 years | 3.1 (1.9, 4.8) | - | 3.1 (1.9, 4.8) | 0.1 (0.1, 0.1) | 0.0 (0.0, 0.0) | 1.3 (0.8, 1.9) | 1.4 (0.9, 2.1) | 4.5 (2.8, 6.8) |

| | | Costs in millions, 2022 USD (95% CI) | | | | | | | |
|---|---|---|---|---|---|---|---|---|---|
| | | **Direct medical costs** | | | **Non-medical costs** | | | | ***Total*** |
| **Condition** | **Age group** | *Hospitalization* | *Post-acute care* | *All medical*[b] | *Missed work* | *QALYs lost*[c] | *YLL due to death* | *All non-medical* | |
| | 2–4 years | 3.5 (2.1, 5.4) | - | 3.5 (2.1, 5.4) | 0.1 (0.1, 0.1) | 0.0 (0.0, 0.1) | 0.7 (0.4, 1.1) | 0.9 (0.5, 1.3) | 4.3 (2.6, 6.6) |
| | 5–17 years | 3.4 (2.1, 5.3) | - | 3.4 (2.1, 5.3) | 0.1 (0.1, 0.1) | 0.0 (0.0, 0.1) | 2.0 (1.3, 3.0) | 2.2 (1.4, 3.2) | 5.6 (3.5, 8.4) |
| | 18–49 years | 16.6 (4.6, 42.1) | - | 16.6 (4.6, 42.1) | 0.8 (0.7, 1.0) | 2.9 (2.5, 3.7) | 51.5 (43.2, 66.0) | 55.2 (46.4, 70.8) | 72.5 (55.4, 103.9) |
| | 50–64 years | 24.2 (6.7, 61.1) | 0.3 (0.1, 0.8) | 24.5 (7.0, 61.5) | 1.1 (0.9, 1.4) | 3.6 (3.0, 4.7) | 94.4 (79.0, 121.1) | 99.1 (83.0, 127.2) | 124.7 (97.3, 173.8) |
| | ≥65 years | 71.9 (19.9, 181.7) | 1.9 (0.5, 4.8) | 74.0 (21.9, 184.0) | 0.9 (0.8, 1.2) | 22.7 (19.1, 29.1) | 374.4 (314.6, 479.5) | 398.0 (334.5, 509.8) | 475.8 (381.0, 641.3) |
| | Total[b] | 127.7 (64.2, 245.1) | 2.2 (0.6, 5.6) | 130.2 (66.4, 247.8) | 3.1 (2.6, 3.9) | 29.4 (24.9, 37.5) | 524.3 (444.4, 668.9) | 556.8 (471.9, 710.4) | 690.5 (566.6, 902.4) |

Abbreviations: PCV – pneumococcal conjugate vaccine; CI – confidence interval; QALY – quality adjusted life year; YLL – years of life lost.

[a] PCV24-additional serotypes (not included in PCV20): 2, 9N, 17F, 20B.

[b] Component values may not sum to totals due to rounding and uncertainty propagation methods.

[c] QALY losses due to episode (no sequelae). Calculated only among survivors.

**Table S11. Years life lost due to pneumococcal disease from PCV24- and PCV31-additional serotypes[a]**

| Condition | Age group | Years of life lost (95% CI)[b] | |
|---|---|---|---|
| | | *PCV24-additional serotypes[a]* | *PCV31-additional serotypes[a]* |
| Non-bacteremic pneumonia | <2 years | 17 (11, 25) | 195 (125, 279) |
| | 2–4 years | 9 (6, 14) | 109 (70, 156) |
| | 5–17 years | 27 (17, 39) | 310 (198, 445) |
| | 18–49 years | 675 (566, 864) | 3,113 (2,807, 3,457) |
| | 50–64 years | 1,236 (1,034, 1,585) | 5,702 (5,119, 6,359) |
| | ≥65 years | 4,900 (4,118, 6,276) | 22,615 (20,434, 25,062) |
| | Total | 6,863 (5,817, 8,756) | 32,055 (29,405, 35,054) |
| Meningitis | <2 years | 2 (1, 6) | 24 (18, 32) |
| | 2–4 years | 3 (1, 7) | 30 (22, 40) |
| | 5–17 years | 12 (10, 15) | 53 (50, 57) |
| | 18–49 years | 54 (46, 68) | 247 (233, 264) |
| | 50–64 years | 82 (70, 104) | 376 (354, 403) |
| | ≥65 years | 118 (101, 150) | 544 (513, 582) |
| | Total | 270 (231, 344) | 1,275 (1,197, 1,367) |
| Bacteremic pneumonia | <2 years | 25 (7, 63) | 262 (104, 349) |
| | 2–4 years | 14 (4, 35) | 146 (108, 195) |
| | 5–17 years | 50 (43, 63) | 230 (217, 246) |
| | 18–49 years | 608 (522, 773) | 2,804 (2,641, 3,000) |
| | 50–64 years | 925 (794, 1,178) | 4,271 (4,022, 4,569 |
| | ≥65 years | 1,338 (1,149, 1,703) | 6,178 (5,818, 6,608) |
| | Total | 2,964 (2,541, 3,769) | 13,896 (13,055, 14,879) |
| Other bacteremia | <2 years | 5 (1, 12) | 49 (36, 66) |
| | 2–4 years | 3 (1, 7) | 28 (20, 37) |
| | 5–17 years | 9 (8, 12) | 43 (41, 46) |
| | 18–49 years | 115 (99, 146) | 530 (499, 567) |
| | 50–64 years | 175 (150, 223) | 807 (760, 864) |
| | ≥65 years | 253 (217, 322) | 1,168 (1,100, 1,249) |
| | Total | 560 (480, 712) | 2,626 (2,467, 2,812) |
| All IPD | <2 years | 32 (9, 80) | 335 (248, 447) |
| | 2–4 years | 19 (6, 49) | 203 (151, 271) |
| | 5–17 years | 71 (61, 90) | 327 (308, 350) |
| | 18–49 years | 776 (666, 987) | 3,582 (3,373, 3831) |
| | 50–64 years | 1,182 (1,015, 1,504) | 5,455 (5,137, 5,835) |
| | ≥65 years | 1,709 (1,467, 2,175) | 7,890 (7,430, 8,440) |
| | Total | 3,794 (3,253, 4,825) | 17,797 (16,719, 19,058) |

Abbreviations: PCV – pneumococcal conjugate vaccine; IPD – invasive pneumococcal disease; CI – confidence interval.

[a] PCV24-additional serotypes (not included in PCV20): 2, 9N, 17F, 20B. PCV31-additional serotypes: 2, 9N, 17F, 20B, 7C, 15A, 16F, 23A, 23B, 31, 35B.

[b] All future values discounted 3% per year.

**Table S12. Annual health economic burden attributable to invasive PCV24-additional serotype[a] pneumococcal disease**

| | | Costs in millions, 2022 USD (95% CI) | | | | | | | | |
|---|---|---|---|---|---|---|---|---|---|---|
| | | Direct medical costs | | | Non-medical costs | | | | | |
| **Condition** | **Age group** | *Hospitali-zation* | *Post-acute care* | *Total medical[b]* | *Missed work* | *QALYs lost, acute[c]* | *YLL due to death[d]* | *QALYs lost, sequelae[d,e]* | *All non-medical[b]* | **Total[b]** |
| Meningitis | <2 years | 0.1 (0.0, 0.2) | 0.1 (0.0, 0.3) | 0.2 (0.1, 0.6) | 0.0 (0.0, 0.0) | 0.0 (0.0, 0.0) | 0.2 (0.0, 0.4) | 0.3 (0.1, 0.6) | 0.4 (0.1, 1.1) | 0.7 (0.2, 1.7) |
| | 2–4 years | 0.1 (0.0, 0.1) | 0.1 (0.0, 0.2) | 0.1 (0.0, 0.3) | 0.0 (0.0, 0.0) | 0.0 (0.0, 0.0) | 0.2 (0.1, 0.5) | 0.1 (0.0, 0.3) | 0.4 (0.1, 0.9) | 0.5 (0.1, 1.2) |
| | 5–17 years | 0.3 (0.2, 0.4) | 0.4 (0.3, 0.5) | 0.6 (0.5, 0.8) | 0.0 (0.0, 0.0) | 0.0 (0.0, 0.0) | 0.9 (0.8, 1.1) | 0.8 (0.7, 1.1) | 1.7 (1.5, 2.2) | 2.4 (2.0, 3.0) |
| | 18–49 years | 0.6 (0.2, 1.4) | 1.0 (0.9, 1.3) | 1.6 (1.1, 2.5) | 0.0 (0.0, 0.0) | 0.1 (0.1, 0.1) | 4.1 (3.5, 5.2) | 8.5 (7.3, 10.8) | 12.7 (10.9, 16.1) | 14.3 (12.2, 18.2) |
| | 50–64 years | 1.2 (0.3, 2.9) | 1.7 (1.4, 2.1) | 2.8 (1.9, 4.7) | 0.0 (0.0, 0.1) | 0.1 (0.1, 0.2) | 6.2 (5.3, 7.9) | 9.7 (8.3, 12.4) | 16.1 (13.9, 20.5) | 19.0 (16.2, 24.4) |
| | ≥65 years | 1.2 (0.3, 2.9) | 2.2 (1.9, 2.8) | 3.4 (2.4, 5.3) | 0.0 (0.0, 0.0) | 0.2 (0.1, 0.2) | 9.0 (7.7, 11.5) | 7.2 (6.2, 9.1) | 16.4 (14.0, 20.8) | 19.8 (16.8, 25.3) |
| | Total[b] | 3.5 (1.9, 6.0) | 5.5 (4.7, 6.9) | 9.0 (7.0, 12.3) | 0.1 (0.1, 0.1) | 0.4 (0.3, 0.5) | 20.6 (17.7, 26.2) | 26.6 (22.8, 33.9) | 47.8 (40.9, 60.7) | 56.8 (48.5, 72.3) |
| Bacteremic pneumonia | <2 years | 0.3 (0.1, 0.8) | - | 0.3 (0.1, 0.8) | 0.0 (0.0, 0.0) | 0.0 (0.0, 0.0) | 1.9 (0.5, 4.8) | - | 1.9 (0.6, 4.8) | 2.3 (0.6, 5.6) |
| | 2–4 years | 0.2 (0.1, 0.6) | - | 0.2 (0.1, 0.6) | 0.0 (0.0, 0.0) | 0.0 (0.0, 0.0) | 1.1 (0.3, 2.7) | - | 1.1 (0.3, 2.7) | 1.3 (0.4, 3.3) |
| | 5–17 years | 0.9 (0.7, 1.2) | - | 0.9 (0.7, 1.2) | 0.0 (0.0, 0.0) | 0.0 (0.0, 0.0) | 3.8 (3.3, 4.9) | - | 3.9 (3.3, 4.9) | 4.8 (4.1, 6.1) |
| | 18–49 years | 6.3 (1.8, 16.0) | - | 6.3 (1.8, 16.0) | 0.4 (0.3, 0.5) | 0.5 (0.5, 0.7) | 46.4 (60.7, 90.0) | - | 47.3 (40.6, 60.2) | 54.0 (44.8, 70.7) |

| | | Costs in millions, 2022 USD (95% CI) | | | | | | | | |
|---|---|---|---|---|---|---|---|---|---|---|
| | | **Direct medical costs** | | | **Non-medical costs** | | | | | |
| **Condition** | **Age group** | *Hospitali-zation* | *Post-acute care* | *Total medical*[b] | *Missed work* | *QALYs lost, acute*[c] | *YLL due to death*[d] | *QALYs lost, sequelae*[d,e] | *All non-medical*[b] | **Total**[b] |
| | 50–64 years | 13.0 (3.6, 32.8) | 9.9 (8.5, 12.7) | 23.1 (13.3, 43.5) | 0.6 (0.5, 0.7) | 0.8 (0.7, 1.1) | 70.7 (60.7, 90.0) | - | 72.1 (61.9, 91.8) | 95.8 (78.7, 126.8) |
| | ≥65 years | 13.2 (3.7, 33.2) | 25.9 (22.3, 33.0) | 39.4 (28.3, 61.4) | 0.2 (0.2, 0.2) | 2.1 (1.8, 2.7) | 102.2 (87.8, 130.1) | - | 104.5 (89.7, 133.0) | 144.5 (121.2, 186.8) |
| | Total[b] | 36.0 (18.5, 64.8) | 35.9 (30.8, 45.6) | 72.2 (52.4, 104.9) | 1.2 (1.0, 1.5) | 3.5 (3.0, 4.5) | 226.4 (194.1, 288.0) | - | 231.1 (198.2, 293.9) | 303.8 (256.8, 388.8) |
| Other bacteremia | <2 years | 0.1 (0.0, 0.2) | - | 0.1 (0.0, 0.2) | 0.0 (0.0, 0.0) | 0.0 (0.0, 0.0) | 0.4 (0.1, 0.9) | - | 0.4 (0.1, 0.9) | 0.4 (0.1, 1.1) |
| | 2–4 years | 0.0 (0.0, 0.1) | - | 0.0 (0.0, 0.1) | 0.0 (0.0, 0.0) | 0.0 (0.0, 0.0) | 0.2 (0.1, 0.5) | - | 0.2 (0.1, 0.5) | 0.2 (0.1, 0.6) |
| | 5–17 years | 0.2 (0.1, 0.2) | - | 0.2 (0.1, 0.2) | 0.0 (0.0, 0.0) | 0.0 (0.0, 0.0) | 0.7 (0.6, 0.9) | - | 0.7 (0.6, 0.9) | 0.9 (0.8, 1.1) |
| | 18–49 years | 1.2 (0.3, 3.0) | - | 1.2 (0.3, 3.0) | 0.1 (0.1, 0.1) | 0.2 (0.1, 0.2) | 8.8 (7.5, 11.2) | - | 9.0 (7.7, 11.5) | 10.3 (8.5, 13.4) |
| | 50–64 years | 2.5 (0.7, 6.2) | - | 2.5 (0.7, 6.2) | 0.1 (0.1, 0.1) | 0.3 (0.2, 0.3) | 13.4 (11.5, 17.0) | - | 13.7 (11.8, 17.5) | 16.3 (13.3, 21.8) |
| | ≥65 years | 2.5 (0.7, 6.3) | - | 2.5 (0.7, 6.3) | 0.0 (0.0, 0.0) | 0.3 (0.3, 0.4) | 19.3 (16.6, 24.6) | - | 19.7 (16.9, 25.0) | 22.3 (18.5, 29.1) |
| | Total[b] | 6.8 (3.5, 12.3) | - | 6.8 (3.5, 12.3) | 0.2 (0.2, 0.3) | 0.7 (0.6, 0.9) | 42.8 (36.7, 54.4) | - | 43.7 (37.5, 55.6) | 50.7 (42.7, 65.0) |
| All[b] | <2 years | 0.5 (0.1, 1.2) | 0.1 (0.0, 0.3) | 0.6 (0.2, 1.6) | 0.0 (0.0, 0.1) | 0.0 (0.0, 0.0) | 2.5 (0.7, 6.1) | 0.3 (0.1, 0.6) | 2.7 (0.8, 6.8) | 3.4 (1.0, 8.4) |

| | | Costs in millions, 2022 USD (95% CI) | | | | | | | | |
|---|---|---|---|---|---|---|---|---|---|---|
| | | Direct medical costs | | | Non-medical costs | | | | | |
| **Condition** | **Age group** | *Hospitali-zation* | *Post-acute care* | *Total medical*[b] | *Missed work* | *QALYs lost, acute*[c] | *YLL due to death*[d] | *QALYs lost, sequelae*[d,e] | *All non-medical*[b] | **Total**[b] |
| | 2–4 years | 0.3 (0.1, 0.9) | 0.1 (0.0, 0.2) | 0.4 (0.1, 1.1) | 0.0 (0.0, 0.0) | 0.0 (0.0, 0.0) | 1.5 (0.4, 3.7) | 0.1 (0.0, 0.3) | 1.6 (0.5, 4.1) | 2.1 (0.6, 5.2) |
| | 5–17 years | 1.3 (1.0, 1.8) | 0.4 (0.3, 0.5) | 1.7 (1.4, 2.2) | 0.0 (0.0, 0.1) | 0.0 (0.0, 0.0) | 5.4 (4.6, 6.9) | 0.8 (0.7, 1.1) | 6.3 (5.4, 8.0) | 8.0 (6.9, 10.2) |
| | 18–49 years | 8.1 (2.2, 20.4) | 1.0 (0.9, 1.3) | 9.1 (3.3, 21.5) | 0.5 (0.4, 0.6) | 0.8 (0.7, 1.0) | 59.3 (50.9, 75.4) | 8.5 (7.3, 10.8) | 69.0 (59.2, 87.8) | 78.6 (65.6, 102.1) |
| | 50–64 years | 16.6 (4.6, 41.9) | 11.6 (10.0, 14.8) | 28.4 (15.9, 54.3) | 0.7 (0.6, 0.9) | 1.2 (1.1, 1.6) | 90.3 (77.5, 114.9) | 9.7 (8.3, 12.4) | 102.0 (87.6, 129.8) | 131.2 (108.3, 172.6) |
| | ≥65 years | 16.9 (4.7, 42.5) | 28.1 (24.1, 35.7) | 45.3 (31.5, 72.9) | 0.2 (0.2, 0.3) | 2.6 (2.2, 3.3) | 130.6 (112.1, 166.2) | 7.2 (6.2, 9.1) | 140.6 (120.7, 178.9) | 186.6 (156.6, 241.1) |
| | Total[b] | 46.3 (23.9, 83.1) | 41.3 (35.5, 52.6) | 88.0 (63.0, 129.2) | 1.5 (1.3, 1.9) | 4.6 (4.0, 5.9) | 289.9 (248.5, 368.6) | 26.6 (22.8, 33.9) | 322.6 (276.6, 410.3) | 411.3 (348.1, 525.9) |

Abbreviations: PCV – pneumococcal conjugate vaccine; CI – confidence interval; QALY – quality adjusted life year; YLL – years of life lost.

[a] PCV24-additional serotypes (not included in PCV20): 2, 9N, 17F, 20B.

[b] Component values may not sum to totals due to rounding.

[c] QALY losses due to episode (no sequelae). Calculated only among survivors.

[d] Future values discounted 3% per year.

[e] QALY losses due to sequelae of meningitis (disability, deafness, hearing loss).

**Table S13. Quality adjusted life years lost due to sequelae of pneumococcal meningitis from PCV24- and PCV31-additional serotypes[a]**

| Meningitis sequelae | Age group | QALYs lost (95% CI)[b] | |
|---|---|---|---|
| | | *PCV24-additional serotypes* [a] | *PCV31-additional serotypes* [a] |
| Disability | <2 years | 2 (0, 4) | 17 (13, 23) |
| | 2–4 years | 1 (0, 2) | 9 (7, 12) |
| | 5–17 years | 7 (6, 8) | 30 (29, 33) |
| | 18–49 years | 85 (73, 108) | 393 (370, 421) |
| | 50–64 years | 98 (84, 125) | 452 (426, 484) |
| | ≥65 years | 72 (62, 92) | 333 (314, 357) |
| | Total | 265 (227, 337) | 1,236 (1,162, 1,323) |
| Deafness | <2 years | 2 (0, 4) | 16 (12, 21) |
| | 2–4 years | 1 (0, 2) | 9 (6, 11) |
| | 5–17 years | 4 (3, 5) | 18 (17, 19) |
| | 18–49 years | 24 (21, 31) | 111 (105, 119) |
| | 50–64 years | 28 (24, 35) | 128 (120, 137) |
| | ≥65 years | 20 (18, 26) | 94 (89, 101) |
| | Total | 79 (67, 100) | 376 (352, 403) |
| Hearing loss | <2 years | 0 (0, 1) | 2 (2, 3) |
| | 2–4 years | 0 (0, 0) | 1 (1, 2) |
| | 5–17 years | 1 (1, 1) | 3 (3, 3) |
| | 18–49 years | 1 (1, 2) | 6 (6, 7) |
| | 50–64 years | 2 (1, 2) | 7 (7, 8) |
| | ≥65 years | 1 (1, 2) | 5 (5, 6) |
| | Total | 5 (4, 7) | 26 (24, 28) |
| All | <2 years | 3 (1, 8) | 35 (26, 47) |
| | 2–4 years | 2 (1, 5) | 19 (14, 26) |
| | 5–17 years | 1 (9, 14) | 51 (48, 54) |
| | 18–49 years | 11 (95, 141) | 511 (481, 547) |
| | 50–64 years | 127 (109, 162) | 587 (553, 628) |
| | ≥65 years | 94 (81, 119) | 433 (408, 463) |
| | Total | 349 (299, 443) | 1,637 (1,538, 1,754) |

Abbreviations: PCV – pneumococcal conjugate vaccine; IPD – invasive pneumococcal disease; CI – confidence interval.

[a] PCV24-additional serotypes (not included in PCV20): 2, 9N, 17F, 20B. PCV31-additional serotypes: 2, 9N, 17F, 20B, 7C, 15A, 16F, 23A, 23B, 31, 35B.

[b] All future values discounted 3% per year.

**Table S14. Annual outpatient health economic burden attributable to non-invasive PCV24-additional serotype[a] pneumococcal disease**

| Condition | Age group | Direct medical costs in millions, 2022 USD (95% CI) | | | | Other costs in millions, 2022 USD (95% CI) | | | | Total costs in millions, 2022 USD (95% CI) |
|---|---|---|---|---|---|---|---|---|---|---|
| | | *Visits[b]* | *Antibiotics* | *Surgery* | *All medical* | *Out of pocket* | *Missed work* | *QALYs lost* | *All non-medical* | |
| AOM | <2 years | 2.0 (0.8, 4.3) | 1.0 (0.3, 2.6) | 11.5 (6.2, 20.3) | 14.6 (8.5, 24.6) | 0.5 (0.1, 1.5) | 3.1 (2.1, 4.6) | 3.2 (2.1, 4.8) | 7.0 (4.6, 10.4) | 21.6 (13.5, 34.2) |
| | 2–4 years | 1.8 (0.7, 4.0) | 1.2 (0.3, 3.3) | 6.9 (2.7, 14.8) | 10.2 (5.2, 19.0) | 0.5 (0.1, 1.4) | 2.9 (1.9, 4.3) | 3.0 (2.0, 4.4) | 6.4 (4.3, 9.5) | 16.7 (10.2, 27.4) |
| | 5–17 years | 1.3 (0.5, 3.0) | 1.3 (0.4, 3.4) | - | 2.7 (1.2, 5.4) | 0.4 (0.1, 1.1) | 2.3 (1.4, 3.5) | 2.3 (1.5, 3.6) | 5.1 (3.1, 7.9) | 7.8 (4.9, 12.4) |
| | Total[c] | 5.1 (2.0, 10.8) | 3.8 (1.8, 7.0) | 18.7 (10.9, 31.1) | 28.0 (17.9, 43.2) | 1.5 (0.4, 3.8) | 8.4 (6.2, 11.3) | 8.6 (6.4, 11.6) | 18.6 (13.7, 25.5) | 46.7 (32.8, 66.9) |
| Sinusitis | 5–17 years | 2.0 (0.6, 5.4) | 2.4 (0.6, 7.0) | 1.3 (0.3, 3.8) | 6.1 (2.5, 13.1) | 0.8 (0.2, 2.2) | 4.4 (2.4, 7.4) | 1.1 (0.6, 1.8) | 6.3 (3.4, 10.8) | 12.6 (6.6, 22.5) |
| | 18–49 years | 8.8 (3.0, 21.7) | 11.1 (2.9, 30.3) | 5.5 (1.5, 15.1) | 26.9 (11.9, 54.1) | 3.2 (0.8, 8.8) | 18.7 (11.4, 27.3) | 4.6 (2.8, 6.7) | 26.8 (16.1, 40.2) | 54.1 (30.4, 89.7) |
| | 50–64 years | 4.3 (1.4, 10.8) | 5.4 (1.4, 14.8) | 2.8 (0.7, 7.7) | 13.2 (5.5, 27.4) | 1.6 (0.4, 4.5) | 9.0 (5.4, 13.1) | 2.3 (1.4, 3.4) | 13.1 (7.8, 19.7) | 26.4 (14.6, 44.9) |
| | ≥65 years | 1.7 (0.5, 4.5) | 2.1 (0.5, 6.0) | 1.2 (0.3, 3.2) | 5.3 (2.3, 10.9) | 0.7 (0.2, 1.9) | 0.8 (0.5, 1.3) | 1.0 (0.6, 1.5) | 2.5 (1.4, 4.3) | 7.8 (4.0, 14.4) |
| | Total[c] | 16.8 (5.6, 42.1) | 22.4 (10.0, 45.9) | 10.9 (2.8, 29.6) | 52.3 (25.2, 98.1) | 6.3 (1.7, 17.2) | 33.0 (20.1, 48.1) | 9.0 (5.5, 13.1) | 48.9 (29.3, 73.5) | 101.7 (58.0, 164.7) |
| Pneumonia | <2 years | 0.2 (0.1, 0.5) | 0.1 (0.0, 0.4) | - | 0.4 (0.2, 0.8) | 0.1 (0.0, 0.2) | 3.2 (1.6, 5.9) | 3.6 (1.8, 6.6) | 6.8 (3.4, 12.6) | 7.2 (3.6, 13.2) |
| | 2–4 years | 0.2 (0.1, 0.5) | 0.2 (0.1, 0.7) | - | 0.5 (0.2, 1.0) | 0.1 (0.0, 0.2) | 3.2 (1.6, 5.8) | 3.6 (1.8, 6.5) | 6.8 (3.4, 12.4) | 7.3 (3.7, 13.2) |

| Condition | Age group | Direct medical costs in millions, 2022 USD (95% CI) | | | | Other costs in millions, 2022 USD (95% CI) | | | | Total costs in millions, 2022 USD (95% CI) |
|---|---|---|---|---|---|---|---|---|---|---|
| | | *Visits*[b] | *Antibiotics* | *Surgery* | *All medical* | *Out of pocket* | *Missed work* | *QALYs lost* | *All non-medical* | |
| | 5–17 years | 0.3 (0.1, 0.8) | 0.2 (0.1, 0.7) | - | 0.6 (0.2, 1.4) | 0.1 (0.0, 0.3) | 5.0 (2.5, 9.2) | 5.6 (2.8, 10.3) | 10.7 (5.3, 19.7) | 11.3 (5.7, 20.7) |
| | 18–49 years | 0.7 (0.3, 1.4) | 0.4 (0.1, 1.2) | - | 1.1 (0.5, 2.2) | 0.1 (0.0, 0.3) | 6.1 (4.5, 8.4) | 2.9 (2.2, 4.0) | 9.2 (6.8, 12.6) | 10.3 (7.7, 14.2) |
| | 50–64 years | 0.3 (0.1, 0.6) | 0.2 (0.1, 0.6) | - | 0.5 (0.3, 1.0) | 0.1 (0.0, 0.1) | 2.6 (1.9, 3.6) | 1.3 (1.0, 1.8) | 4.0 (3.0, 5.5) | 4.6 (3.4, 6.3) |
| | ≥65 years | 0.7 (0.3, 1.5) | 0.4 (0.1, 1.2) | - | 1.2 (0.6, 2.2) | 0.1 (0.0, 0.4) | 1.6 (1.1, 2.3) | 16.6 (11.6, 23.9) | 18.4 (12.9, 26.4) | 19.6 (13.8, 28.0) |
| | Total[c] | 2.5 (1.2, 4.6) | 1.9 (1.0, 3.3) | - | 4.4 (2.6, 7.2) | 0.5 (0.1, 1.3) | 22.1 (16.2, 30.2) | 34.1 (25.6, 45.6) | 56.8 (42.8, 76.0) | 61.3 (46.3, 81.8) |
| All[c] | <2 years | 2.2 (0.9, 4.7) | 1.1 (0.4, 2.8) | 11.5 (6.2, 20.3) | 15.0 (8.9, 25.0) | 0.6 (0.2 1.6) | 6.4 (4.3, 9.5) | 6.9 (4.6, 10.3) | 13.9 (9.4, 20.7) | 29.2 (19.6, 43.1) |
| | 2–4 years | 2.0 (0.8, 4.4) | 1.5 (0.5, 3.6) | 6.9 (2.7, 14.8) | 10.7 (5.7, 19.5) | 0.6 (0.2, 1.5) | 6.1 (4.1, 9.1) | 6.6 (4.4, 9.9) | 13.4 (9.0, 19.9) | 24.3 (16.2, 36.5) |
| | 5–17 years | 3.7 (1.4, 8.6) | 4.2 (1.7, 9.3) | 1.3 (0.3, 3.8) | 9.6 (4.7, 18.1) | 1.3 (0.3, 3.4) | 11.9 (8.1, 17.2) | 9.1 (5.9, 14.1) | 22.4 (15.2, 33.0) | 32.3 (21.8, 47.4) |
| | 18–49 years | 9.5 (3.5, 22.5) | 11.6 (3.4, 30.9) | 5.5 (1.5, 15.1) | 28.1 (12.9, 55.5) | 3.3 (0.9, 9.1) | 24.9 (17.1, 34.0) | 7.6 (5.5, 10.0) | 36.1 (24.7, 50.3) | 64.5 (40.2, 101.0) |
| | 50–64 years | 4.6 (1.6, 11.2) | 5.7 (1.7, 15.1) | 2.8 (0.7, 7.7) | 13.8 (6.0, 28.0) | 1.7 (0.4, 4.6) | 11.6 (7.9, 16.0) | 3.7 (2.6, 4.9) | 17.1 (11.6, 24.1) | 31.1 (19.0, 49.9) |
| | ≥65 years | 2.5 (1.1, 5.3) | 2.6 (0.8, 6.7) | 1.2 (0.3, 3.2) | 6.5 (3.2, 12.4) | 0.8 (0.2, 2.2) | 2.5 (1.8, 3.3) | 17.6 (12.5, 24.9) | 21.0 (15.2, 29.3) | 27.7 (20.0, 38.6) |

| Condition | Age group | Direct medical costs in millions, 2022 USD (95% CI) | | | | Other costs in millions, 2022 USD (95% CI) | | | | Total costs in millions, 2022 USD (95% CI) |
|---|---|---|---|---|---|---|---|---|---|---|
| | | *Visits*[b] | *Antibiotics* | *Surgery* | *All medical* | *Out of pocket* | *Missed work* | *QALYs lost* | *All non-medical* | |
| | Total[c] | 24.6 (9.8, 55.3) | 28.3 (14.9, 52.6) | 30.4 (17.6, 52.1) | 85.3 (52.0, 138.6) | 8.4 (2.2, 22.0) | 63.8 (48.0, 82.6) | 51.9 (41.3, 65.5) | 124.8 (96.6, 160.5) | 210.9 (155.3, 287.9) |

Abbreviations: PCV – pneumococcal conjugate vaccine; CI – confidence interval; AOM – acute otitis media; QALYs – quality adjusted life years.

[a] PCV24-additional serotypes (not included in PCV20): 2, 9N, 17F, 20B.

[b] Includes both office and emergency department visits, weighted as detailed in **Table 1**.

[c] Component values may not sum to totals due to rounding.

**Table S15. Annual inpatient health economic burden attributable to non-invasive PCV31-additional serotype[a] pneumococcal disease**

| | | Costs in millions, 2022 USD (95% CI) | | | | | | | |
|---|---|---|---|---|---|---|---|---|---|
| | | Direct medical costs | | | Non-medical costs | | | | ***Total*** |
| **Condition** | **Age group** | *Hospitalization* | *Post-acute care* | *All medical[b]* | *Missed work* | *QALYs lost[c]* | *YLL due to death* | *All non-medical* | |
| Mastoiditis | <2 years | 0.4 (0.1, 1.2) | - | 0.4 (0.1, 1.2) | 0.0 (0.0, 0.0) | 0.0 (0.0, 0.0) | - | 0.0 (0.0, 0.0) | 0.4 (0.1, 1.2) |
| | 2–4 years | 0.3 (0.1, 0.8) | - | 0.3 (0.1, 0.8) | 0.0 (0.0, 0.0) | 0.0 (0.0, 0.0) | - | 0.0 (0.0, 0.0) | 0.3 (0.1, 0.9) |
| | 5–17 years | 1.3 (0.3, 3.7) | - | 1.3 (0.3, 3.7) | 0.1 (0.0, 0.1) | 0.0 (0.0, 0.0) | - | 0.1 (0.0, 0.1) | 1.4 (0.4, 3.8) |
| | Total[b] | 2.0 (0.5, 5.7) | - | 2.0 (0.5, 5.7) | 0.1 (0.1, 0.2) | 0.0 (0.0, 0.0) | - | 0.1 (0.1, 0.2) | 2.2 (0.6, 5.9) |
| Pneumonia | <2 years | 36.0 (22.1, 54.4) | - | 36.0 (22.1, 54.4) | 1.0 (0.7, 1.5) | 0.4 (0.3, 0.6) | 14.9 (9.5, 21.3) | 16.4 (10.5, 23.4) | 52.4 (32.9, 77.1) |
| | 2–4 years | 40.5 (24.9, 61.3) | - | 40.5 (24.9, 61.3) | 1.2 (0.7, 1.7) | 0.4 (0.3, 0.6) | 8.3 (5.3, 11.9) | 9.9 (6.4, 14.2) | 50.5 (31.5, 75.1) |
| | 5–17 years | 39.6 (24.3, 60.0) | - | 39.6 (24.3, 60.0) | 1.1 (0.7, 1.6) | 0.4 (0.3, 0.6) | 23.7 (15.1, 34.0) | 25.3 (16.1, 36.2) | 65.0 (40.9, 95.3) |
| | 18–49 years | 76.3 (21.3, 189.0) | - | 76.3 (21.3, 189.0) | 3.6 (3.2, 4.0) | 13.5 (12.2, 15.0) | 237.8 (214.5, 264.1) | 254.9 (229.9, 283.1) | 332.1 (267.9, 450.4) |
| | 50–64 years | 110.8 (31.0, 274.4) | 1.5 (0.4, 3.7) | 112.4 (32.5, 276.2) | 5.1 (4.5, 5.6) | 16.8 (15.1, 18.7) | 435.6 (391.1, 485.8) | 457.5 (410.7, 510.2) | 571.7 (471.7, 747.3) |
| | ≥65 years | 329.8 (92.1, 816.6) | 8.7 (2.4, 21.7) | 339.5 (101.4, 826.4) | 4.3 (3.9, 4.8) | 104.8 (94.7, 116.1) | 1,727.8 (1,561.1, 1,914.7) | 1,836.8 (1,659.7, 2,035.6) | 2,184.1 (1,859.3, 2,719.8) |
| | Total[b] | 657.6 (366.2, 1167.5) | 10.2 (2.9, 25.3) | 668.9 (376.8, 1179.3) | 16.3 (14.6, 18.2) | 136.4 (124.8, 149.5) | 2,449.0 (2,246.5, 2,678.1) | 2,601.7 (2,386.6, 2,845.1) | 3,278.8 (2,878.6, 3,861.9) |

| | | Costs in millions, 2022 USD (95% CI) | | | | | | | |
|---|---|---|---|---|---|---|---|---|---|
| | | **Direct medical costs** | | | **Non-medical costs** | | | | ***Total*** |
| **Condition** | **Age group** | *Hospitalization* | *Post-acute care* | *All medical[b]* | *Missed work* | *QALYs lost[c]* | *YLL due to death* | *All non-medical* | |
| All[b] | <2 years | 36.5 (22.6, 54.9) | - | 36.5 (22.6, 54.9) | 1.1 (0.7, 1.5) | 0.4 (0.3, 0.6) | 14.9 (9.5, 21.3) | 16.4 (10.5, 23.4) | 52.9 (33.4, 77.6) |
| | 2–4 years | 40.9 (25.2, 61.7) | - | 40.9 (25.2, 61.7) | 1.2 (0.8, 1.7) | 0.4 (0.3, 0.6) | 8.3 (5.3, 11.9) | 10.0 (6.4, 14.3) | 50.9 (31.8, 75.4) |
| | 5–17 years | 41.2 (25.7, 61.6) | - | 41.2 (25.7, 61.6) | 1.2 (0.8, 1.7) | 0.4 (0.3, 0.6) | 23.7 (15.1, 34.0) | 25.4 (16.2, 36.3) | 66.6 (42.4, 96.9) |
| | 18–49 years | 76.3 (21.3, 189.0) | - | 76.3 (21.3, 189.0) | 3.6 (3.2, 4.0) | 13.5 (12.2, 15.0) | 237.8 (214.5, 264.1) | 254.9 (229.9, 283.1) | 332.1 (267.9, 450.4) |
| | 50–64 years | 110.8 (31.0, 274.4) | 1.5 (0.4, 3.7) | 112.4 (32.5, 276.2) | 5.1 (4.5, 5.6) | 16.8 (15.1, 18.7) | 435.6 (391.1, 485.8) | 457.5 (410.7, 510.2) | 571.7 (471.7, 747.3) |
| | ≥65 years | 329.8 (92.1, 816.6) | 8.7 (2.4, 21.7) | 339.5 (101.4, 826.4) | 4.3 (3.9, 4.8) | 104.8 (94.7, 116.1) | 1,727.8 (1,561.1, 1,914.7) | 1836.8 (1659.7, 2035.6) | 2,184.1 (1,859.3, 2,719.8) |
| | Total[b] | 659.9 (368.4, 1,169.8) | 10.2 (2.9, 25.3) | 671.2 (379.0, 1,181.6) | 16.4 (14.8, 18.3) | 136.4 (124.8, 149.5) | 2,449.0 (2,246.5, 2,678.1) | 2601.8 (2386.7, 2845.2) | 3,281.3 (2,880.9 3,864.5) |

Abbreviations: PCV – pneumococcal conjugate vaccine; CI – confidence interval; QALY – quality adjusted life year; YLL – years of life lost.

[a] PCV31-additional serotypes: 2, 9N, 17F, 20B, 7C, 15A, 16F, 23A, 23B, 31, 35B.

**[b]** Component values may not sum exactly to totals due to rounding.

[c] QALY losses due to episode (no sequelae). Calculated only among survivors.

**Table S16. Annual health economic burden attributable to invasive PCV31-additional serotype[a] pneumococcal disease**

| | | Costs in millions, 2022 USD (95% CI) | | | | | | | | |
|---|---|---|---|---|---|---|---|---|---|---|
| | | Direct medical costs | | | Non-medical costs | | | | | |
| **Condition** | **Age group** | *Hospitali-zation* | *Post-acute care* | *Total medical*[b] | *Missed work* | *QALYs lost, acute*[c] | *YLL due to death*[d] | *QALYs lost, sequelae*[d,e] | *All non-medical*[b] | **Total**[b] |
| Meningitis | <2 years | 1.0 (0.7, 1.5) | 1.4 (1.0, 1.9) | 2.4 (1.8, 3.3) | 0.1 (0.0, 0.1) | 0.0 (0.0, 0.0) | 1.8 (1.3, 2.4) | 2.7 (2.0, 3.6) | 4.6 (3.4, 6.1) | 7.0 (5.2, 9.4) |
| | 2–4 years | 0.6 (0.4, 0.8) | 0.8 (0.6, 1.1) | 1.4 (1.0, 1.9) | 0.0 (0.0, 0.0) | 0.0 (0.0, 0.0) | 2.3 (1.7, 3.0) | 1.5 (1.1, 2.0) | 3.8 (2.8, 5.0) | 5.1 (3.8, 6.8) |
| | 5–17 years | 1.2 (1.0, 1.5) | 1.7 (1.6, 1.8) | 2.9 (2.7, 3.3) | 0.0 (0.0, 0.0) | 0.0 (0.0, 0.0) | 4.1 (3.8, 4.4) | 3.9 (3.7, 4.2) | 8.0 (7.5, 8.6) | 10.9 (10.3, 11.8) |
| | 18–49 years | 2.6 (0.7, 6.3) | 4.7 (4.5, 5.1) | 7.3 (5.4, 11.1) | 0.1 (0.1, 0.1) | 0.4 (0.4, 0.4) | 18.9 (17.8, 20.2) | 39.0 (36.8, 41.8) | 58.4 (55.0, 62.5) | 65.9 (61.4, 71.6) |
| | 50–64 years | 5.3 (1.5, 13.0) | 7.7 (7.2, 8.2) | 13.0 (9.1, 20.7) | 0.2 (0.2, 0.2) | 0.6 (0.6, 0.7) | 28.8 (27.1, 30.8) | 44.9 (42.3, 48.0) | 74.5 (70.1, 79.7) | 87.7 (81.0, 97.2) |
| | ≥65 years | 5.3 (1.5, 13.2) | 10.0 (9.4, 10.7) | 15.4 (11.4, 23.3) | 0.1 (0.1, 0.1) | 0.8 (0.7, 0.8) | 41.6 (39.2, 44.5) | 33.1 (31.2, 35.4) | 75.5 (71.1, 80.8) | 91.1 (84.3, 100.9) |
| | Total[b] | 16.8 (9.8, 27.7) | 26.4 (24.6, 28.4) | 43.2 (35.7, 54.5) | 0.5 (0.4, 0.5) | 1.8 (1.7, 1.9) | 97.4 (91.4, 104.4) | 125.1 (117.5, 134.0) | 224.8 (211.1, 240.8) | 268.4 (250.1, 290.1) |
| Bacteremic pneumonia | <2 years | 3.4 (2.4, 4.8) | - | 3.4 (2.4, 4.8) | 0.2 (0.1, 0.2) | 0.1 (0.0, 0.1) | 20.0 (14.8, 26.7) | - | 20.2 (15.0, 27.0) | 23.6 (17.5, 31.5) |
| | 2–4 years | 2.5 (1.8, 3.6) | - | 2.5 (1.8, 3.6) | 0.1 (0.1, 0.1) | 0.0 (0.0, 0.0) | 11.2 (8.3, 14.9) | - | 11.3 (8.4, 15.0) | 13.8 (10.2, 18.5) |
| | 5–17 years | 4.2 (3.3, 5.1) | - | 4.2 (3.3, 5.1) | 0.1 (0.1, 0.2) | 0.1 (0.0, 0.1) | 17.6 (16.6, 18.8) | - | 17.8 (16.7, 19.0) | 22.0 (20.5, 23.7) |

| | | Costs in millions, 2022 USD (95% CI) | | | | | | | | |
|---|---|---|---|---|---|---|---|---|---|---|
| | | **Direct medical costs** | | | **Non-medical costs** | | | | | |
| **Condition** | **Age group** | *Hospitali-zation* | *Post-acute care* | *Total medical*[b] | *Missed work* | *QALYs lost, acute*[c] | *YLL due to death*[d] | *QALYs lost, sequelae*[d,e] | *All non-medical*[b] | **Total**[b] |
| | 18–49 years | 29.1 (8.1, 71.6) | - | 29.1 (8.1, 71.6) | 1.8 (1.7, 1.9) | 2.4 (2.3, 2.6) | 214.3 (201.8, 229.2) | - | 218.5 (205.7, 233.7) | 248.2 (22.0, 293.5) |
| | 50–64 years | 59.8 (16.7, 147.4) | 45.9 (43.2, 49.1) | 105.8 (62.5, 193.7) | 2.7 (2.6, 2.9) | 3.9 (3.6, 4.1) | 326.3 (307.3, 349.1) | - | 332.9 (313.5, 356.1) | 439.7 (388.4, 531.5) |
| | ≥65 years | 60.6 (17.0, 149.3) | 119.6 (112.7, 128.0) | 180.4 (135.7, 270.0) | 0.8 (0.8, 0.9) | 9.8 (9.2, 10.4) | 472.0 (444.4, 504.9) | - | 482.6 (454.4, 516.2) | 664.7 (604.0, 762.6) |
| | Total[b] | 168.5 (89.5, 291.1) | 165.6 (155.9, 177.1) | 334.3 (253.6, 458.6) | 5.7 (5.3, 6.1) | 16.2 (15.2, 17.3) | 1061.6 (997.4, 1136.8) | - | 1083.5 (1018.0, 1160.2) | 1420.4 (1301.5, 1575.3) |
| Other bacteremia | <2 years | 0.6 (0.4, 0.9) | - | 0.6 (0.4, 0.9) | 0.0 (0.0, 0.0) | 0.0 (0.0, 0.0) | 3.8 (2.8, 5.0) | - | 3.8 (2.8, 5.1) | 4.4 (3.3, 5.9) |
| | 2–4 years | 0.5 (0.3, 0.7) | - | 0.5 (0.3, 0.7) | 0.0 (0.0, 0.0) | 0.0 (0.0, 0.0) | 2.1 (1.6, 2.8) | - | 2.1 (1.6, 2.8) | 2.6 (1.9, 3.5) |
| | 5–17 years | 0.8 (0.6, 1.0) | - | 0.8 (0.6, 1.0) | 0.0 (0.0, 0.0) | 0.0 (0.0, 0.0) | 3.3 (3.1, 3.6) | - | 3.4 (3.2, 3.6) | 4.1 (3.9, 4.5) |
| | 18–49 years | 5.5 (1.5, 13.5) | - | 5.5 (1.5, 13.5) | 0.3 (0.3, 0.4) | 0.7 (0.7, 0.8) | 40.5 (38.1, 43.3) | - | 41.6 (39.1, 44.5) | 47.2 (42.2, 55.8) |
| | 50–64 years | 11.3 (3.2, 27.9) | - | 11.3 (3.2, 27.9) | 0.5 (0.5, 0,5) | 1.2 (1.1, 1.3) | 61.7 (58.1, 66.0) | - | 63.4 (59.7, 67.8) | 74.8 (65.4, 92.1) |
| | ≥65 years | 11.4 (3.2, 28.2) | - | 11.4 (3.2, 28.2) | 0.2 (0.1, 0.2) | 1.4 (1.3, 1.5) | 89.2 (84.0, 95.4) | - | 90.8 (85.5, 97.1) | 102.5 (92.0, 120.4) |
| | Total[b] | 31.8 (16.9, 55.0) | - | 31.8 (16.9, 55.0) | 1.1 (1.0, 1.2) | 3.3 (3.1, 3.6) | 200.6 (188.5, 214.8) | - | 205.0 (192.6, 219.5) | 237.4 (216.2, 265.4) |

| | | Costs in millions, 2022 USD (95% CI) | | | | | | | | |
|---|---|---|---|---|---|---|---|---|---|---|
| | | Direct medical costs | | | Non-medical costs | | | | | |
| **Condition** | **Age group** | *Hospitali-zation* | *Post-acute care* | *Total medical*[b] | *Missed work* | *QALYs lost, acute*[c] | *YLL due to death*[d] | *QALYs lost, sequelae*[d,e] | *All non-medical*[b] | **Total**[b] |
| All[b] | <2 years | 5.1 (3.6, 7.0) | 1.4 (1.0, 1.9) | 6.5 (4.7, 8.9) | 0.2 (0.2, 0.3) | 0.1 (0.1, 0.1) | 25.6 (18.9, 34.1) | 2.7 (2.0, 3.6) | 28.6 (21.2, 38.1) | 35.1 (25.9, 46.8) |
| | 2–4 years | 3.6 (2.5, 5.1) | 0.8 (0.6, 1.1) | 4.4 (3.1, 6.1) | 0.1 (0,1, 0,2) | 0.0 (0.0, 0.1) | 15.5 (11.5, 20.7) | 1.5 (1.1, 2.0) | 17.2 (12.7, 22.9) | 21.6 (15.9, 28.8) |
| | 5–17 years | 6.2 (5.0, 7.6) | 1.7 (1.6, 1.8) | 7.9 (6.6, 9.3) | 0.2 (0.2, 0.2) | 0.1 (0.1, 0.1) | 25.0 (23.5, 26.7) | 3.9 (3.7, 4.2) | 29.1 (27.4, 31.2) | 37.0 (34.6, 39.9) |
| | 18–49 years | 37.1 (10.4, 91.5) | 4.7 (4.5, 5.1) | 41.9 (15.1, 96.3) | 2.2 (2.1, 2.4) | 3.6 (3.4, 3.8) | 273.6 (257.7, 292.7) | 39.0 (36.8, 41.8) | 318.4 (299.9, 340.6) | 361.3 (326.0, 420.1) |
| | 50–64 years | 76.4 (21.4, 188.3) | 53.6 (50.5, 57.3) | 130.1 (74.8, 242.3) | 3.4 (3.2, 3.7) | 5.7 (5.3, 6.1) | 416.8 (392.5, 445.8) | 44.9 (42.3, 48.0) | 470.8 (443.3, 503.6) | 602.3 (535.3 720.3) |
| | ≥65 years | 77.4 (21.7, 190.7) | 129.6 (122.1, 138.7) | 207.2 (150.5, 321.4) | 1.0 (1.0, 1.1) | 11.9 (11.2, 12.7) | 602.8 (567.6, 644.8) | 33.1 (31.2, 35.4) | 648.8 (611.0, 694.0) | 858.4 (760.4, 983.6) |
| | Total[b] | 217.1 (116.3, 373.7) | 191.9 (180.6, 205.4) | 409.3 (306.4, 567.9) | 7.2 (6.8, 7.8) | 21.3 (20.1, 22.8) | 1359.7 (1277.3, 1456.0) | 125.1 (117.5, 134.0) | 1513.3 (1421.7, 1620.5) | 1926.3 (1768.8, 2129.2) |

Abbreviations: PCV – pneumococcal conjugate vaccine; CI – confidence interval; QALY – quality adjusted life year; YLL – years of life lost.

[a] PCV31-additional serotypes: 2, 9N, 17F, 20B, 7C, 15A, 16F, 23A, 23B, 31, 35B.

**[b]** Component values may not sum to totals due to rounding.

[c] QALY losses due to episode (no sequelae). Calculated only among survivors.

[d] Future values discounted 3% per year.

[e] QALY losses due to sequelae of meningitis (disability, deafness, hearing loss).

**Table S17. Annual outpatient health economic burden attributable to non-invasive PCV31-additional serotype[a] pneumococcal disease**

| Condition | Age group | Direct medical costs in millions, 2022 USD (95% CI) | | | | Other costs in millions, 2022 USD (95% CI) | | | | Total costs in millions, 2022 USD (95% CI) |
|---|---|---|---|---|---|---|---|---|---|---|
| | | *Visits[b]* | *Antibiotics* | *Surgery* | *All medical* | *Out of pocket* | *Missed work* | *QALYs lost* | *All non-medical* | |
| AOM | <2 years | 24.0 (9.5, 51.9) | 11.7 (3.1, 31.6) | 140.8 (78.3, 242.4) | 179.5 (107.1, 292.4) | 6.7 (1.8, 17.9) | 38.6 (26.3, 54.7) | 39.6 (27.0, 56.2) | 85.5 (57.8, 122.8) | 265.6 (171.0, 405.4) |
| | 2–4 years | 22.2 (8.8, 47.7) | 15.1 (4.1, 39.7) | 84.2 (34.2, 177.9) | 124.9 (65.7, 227.2) | 6.2 (1.7, 16.5) | 35.7 (24.7, 50.1) | 36.6 (25.3, 51.4) | 79.0 (54.2, 112.3) | 204.7 (128.8, 326.2) |
| | 5–17 years | 15.6 (5.9, 35.8) | 16.2 (4.4, 41.4) | - | 33.1 (14.9, 65.0) | 4.9 (1.3, 13.3) | 28.1 (18.0, 41.8) | 28.8 (18.5, 42.9) | 62.3 (39.7, 93.6) | 96.3 (61.4, 146.6) |
| | Total[c] | 62.5 (25.5, 129.5) | 46.1 (22.7, 84.0) | 229.3 (137.4, 370.6) | 343.5 (226.8, 511.7) | 18.1 (5.0, 46.1) | 103.1 (80.1, 131.1) | 105.8 (82.2, 134.6) | 228.4 (175.4, 295.9) | 573.1 (418.8, 783.8) |
| Sinusitis | 5–17 years | 23.4 (7.3, 62.4) | 28.1 (7.1, 80.7) | 15.3 (3.9, 44.0) | 71.3 (29.6, 150.3) | 8.9 (2.3, 25.6) | 51.7 (28.8, 84.1) | 12.7 (7.1, 20.7) | 74.1 (40.8, 123.1) | 146.9 (77.7, 257.5) |
| | 18–49 years | 102.2 (34.8, 250.7) | 129.6 (34.1, 350.5) | 64.7 (17.0, 174.7) | 314.1 (140.1, 621.7) | 37.6 (9.9, 101.5) | 218.5 (134.8, 309.3) | 53.7 (33.2 76.1) | 312.9 (190.7, 457.1) | 631.1 (359.6, 1026.4) |
| | 50–64 years | 49.9 (16.3, 125.0) | 63.3 (16.6, 171.5) | 32.8 (8.6, 88.4) | 154.2 (65.2, 314.9) | 19.0 (5.0, 51.4) | 104.7 (64.6, 148.4) | 27.2 (16.8 38.5) | 152.5 (92.7, 223.6) | 308.4 (172.4, 514.3) |
| | ≥65 years | 19.7 (6.0, 51.8) | 24.5 (6.3, 69.1) | 13.4 (3.5, 37.1) | 61.5 (26.6, 125.1) | 7.8 (2.0, 21.6) | 9.9 (5.9, 14.9) | 11.1 (6.6, 16.7) | 29.2 (16.5, 49.2) | 91.5 (47.1, 165.5) |
| | Total[c] | 195.8 (65.3, 485.4) | 261.4 (117.8, 527.6) | 126.6 (33.4, 341.5) | 609.7 (298.1, 1125.5) | 73.6 (19.4, 198.5) | 386.1 (238.5, 545.1) | 105.2 (65.0, 148.3) | 570.6 (347.5, 835.3) | 1187.2 (686.6, 1881.2) |

| Condition | Age group | Direct medical costs in millions, 2022 USD (95% CI) | | | | Other costs in millions, 2022 USD (95% CI) | | | | Total costs in millions, 2022 USD (95% CI) |
|---|---|---|---|---|---|---|---|---|---|---|
| | | *Visits*[b] | *Antibiotics* | *Surgery* | *All medical* | *Out of pocket* | *Missed work* | *QALYs lost* | *All non-medical* | |
| Pneumonia | <2 years | 2.5 (0.9, 6.2) | 1.7 (0.4, 5.0) | - | 4.4 (1.9, 9.4) | 0.7 (0.2, 2.1) | 37.2 (18.8, 67.5) | 41.6 (21.1, 75.5) | 79.6 (40.3, 144.4) | 84.2 (43.1, 151.8) |
| | 2–4 years | 2.5 (0.9, 6.2) | 2.8 (0.7, 7.8) | - | 5.5 (2.3, 11.8) | 0.7 (0.2, 2.0) | 37.1 (19.0, 66.8) | 41.5 (21.2, 74.7) | 79.4 (40.6, 142.8) | 85.2 (44.2, 151.8) |
| | 5–17 years | 3.8 (1.3, 9.6) | 2.9 (0.7, 8.1) | - | 6.9 (2.7, 15.6) | 1.1 (0.3, 3.2) | 58.2 (29.5, 105.6) | 65.1 (33.0, 118.1) | 124.5 (63.0, 225.9) | 131.8 (67.4, 237.5) |
| | 18–49 years | 3.0 (1.3, 6.1) | 2.0 (0.5, 5.3) | - | 5.2 (2.5, 9.7) | 0.5 (0.1, 1.3) | 28.0 (21.6, 35.8) | 13.5 (10.4, 17.2) | 42.1 (32.5, 53.8) | 47.5 (36.6, 60.8) |
| | 50–64 years | 1.3 (0.5, 2.6) | 1.1 (0.3, 2.8) | - | 2.4 (1.2, 4.6) | 0.2 (0.1, 0.6) | 12.1 (9.3, 15.4) | 6.1 (4.7, 7.8) | 18.5 (14.3, 23.6) | 21.0 (16.2, 26.9) |
| | ≥65 years | 3.2 (1.3, 6.8) | 1.9 (0.5, 5.2) | - | 5.3 (2.6, 10.0) | 0.6 (0.2, 1.6) | 7.4 (5.4, 10.0) | 76.5 (55.2, 103.2) | 84.6 (61.1, 114.1) | 90.1 (65.4, 121.2) |
| | Total[c] | 16.8 (8.1, 31.9) | 13.6 (7.5, 23.2) | - | 30.8 (18.3, 50.2) | 3.9 (1.1, 10.2) | 182.9 (126.2, 262.5 | 248.2 (180.8, 339.9) | 435.3 (311.7, 607.1) | 466.7 (335.8, 647.9) |
| All[c] | <2 years | 26.6 (10.6, 56.8) | 13.8 (4.8, 33.8) | 140.8 (78.3, 242.4) | 184.3 (111.2, 297.8) | 7.5 (2.0, 19.6) | 76.6 (53.1, 109.9) | 82.0 (56.5, 118.7) | 166.9 (115.6, 239.4) | 353.6 (244.6, 505.2) |
| | 2–4 years | 24.8 (10.0, 52.7) | 18.4 (6.7, 43.2) | 84.2 (34.2, 177.9) | 130.9 (70.9, 233.6) | 6.9 (1.9, 18.2) | 73.5 (51.1, 105.5) | 78.8 (54.4, 114.3) | 160.0 (111.1, 229.7) | 293.8 (201.2, 427.8) |
| | 5–17 years | 43.6 (16.2, 101.1) | 49.7 (20.4, 108.1) | 15.3 (3.9, 44.0) | 113.4 (56.2, 209.9) | 15.2 (4.1, 40.1) | 140.1 (98.6, 196.5) | 107.7 (72.2, 162.3) | 264.9 (184.8, 378.4) | 381.8 (265.3, 543.6) |

| Condition | Age group | Direct medical costs in millions, 2022 USD (95% CI) | | | | Other costs in millions, 2022 USD (95% CI) | | | | Total costs in millions, 2022 USD (95% CI) |
|---|---|---|---|---|---|---|---|---|---|---|
| | | *Visits*[b] | *Antibiotics* | *Surgery* | *All medical* | *Out of pocket* | *Missed work* | *QALYs lost* | *All non-medical* | |
| | 18–49 years | 105.4 (37.5, 254.2) | 131.8 (36.3, 352.7) | 64.7 (17.0, 174.7) | 319.5 (144.7, 628.0) | 38.2 (10.1, 102.7) | 246.7 (162.6, 337.9) | 67.3 (46.4, 90.0) | 355.2 (232.3, 500.3) | 678.9 (406.4, 1075.4) |
| | 50–64 years | 51.3 (17.5, 126.5) | 64.5 (17.8, 172.7) | 32.8 (8.6, 88.4) | 156.7 (67.4, 317.7) | 19.3 (5.1, 52.0) | 116.9 (76.6, 160.8) | 33.4 (22.8, 44.8) | 171.1 (111.1, 242.5) | 329.5 (193.2, 535.9) |
| | ≥65 years | 23.1 (8.9, 55.4) | 26.6 (7.7, 72.2) | 13.4 (3.5, 37.1) | 67.0 (31.1, 131.9) | 8.5 (2.2, 23.0) | 17.4 (12.8, 23.0) | 87.8 (65.8, 115.0) | 114.6 (86.5, 149.5) | 182.9 (129.0, 262.7) |
| | Total[c] | 277.0 (106.4, 628.8) | 323.6 (170.4, 597.7) | 365.8 (217.7, 609.4) | 990.6 (615.1, 1578.5) | 96.2 (26.0, 250.8) | 675.3 (511.4, 853.3) | 461.0 (375.6, 565.1) | 1241.0 (965.4, 1568.7) | 2239.6 (1656.7, 3026.0) |

Abbreviations: PCV – pneumococcal conjugate vaccine; CI – confidence interval; AOM – acute otitis media; QALYs – quality adjusted life years.

[a] PCV31-additional serotypes: 2, 9N, 17F, 20B, 7C, 15A, 16F, 23A, 23B, 31, 35B.

**[b]** Includes both office and emergency department visits, weighted as detailed in **Table 1**.

[c] Component values may not sum exactly to totals due to rounding.
